# Deleterious, protein-altering variants in the X-linked transcriptional coregulator *ZMYM3* in 22 individuals with a neurodevelopmental delay phenotype

**DOI:** 10.1101/2022.09.29.22279724

**Authors:** Susan M. Hiatt, Slavica Trajkova, Matteo Rossi Sebastiano, E. Christopher Partridge, Fatima E. Abidi, Ashlyn Anderson, Muhammad Ansar, Stylianos E. Antonarakis, Azadeh Azadi, Ruxandra Bachmann-Gagescu, Andrea Bartuli, Caroline Benech, Jennifer L. Berkowitz, Michael J. Betti, Alfredo Brusco, Ashley Cannon, Giulia Caron, Yanmin Chen, Molly M. Crenshaw, Laurence Cuisset, Cynthia J. Curry, Hossein Darvish, Serwet Demirdas, Maria Descartes, Jessica Douglas, David A. Dyment, Houda Zghal Elloumi, Giuseppe Ermondi, Marie Faoucher, Emily G. Farrow, Stephanie A. Felker, Heather Fisher, Anna C. E. Hurst, Pascal Joset, Stanislav Kmoch, Benjamin R. Leadem, Marina Macchiaiolo, Martin Magner, Giorgia Mandrile, Francesca Mattioli, Megan McEown, Sarah K. Meadows, Livija Medne, Naomi J. L. Meeks, Sarah Montgomery, Melanie P. Napier, Marvin Natowicz, Kimberly M. Newberry, Marcello Niceta, Lenka Noskova, Catherine Nowak, Amanda G. Noyes, Matthew Osmond, Verdiana Pullano, Chloé Quélin, Simin Rahimi-Aliabadi, Anita Rauch, Sylvia Redon, Alexandre Reymond, Caitlin R. Schwager, Elizabeth A. Sellars, Angela Scheuerle, Elena Shukarova-Angelovska, Cara Skraban, Bonnie R. Sullivan, Marco Tartaglia, Isabelle Thiffault, Kevin Uguen, Luis A. Umaña, Yolande van Bever, Saskia N. van der Crabben, Marjon A. van Slegtenhorst, Quinten Waisfisz, Richard M. Myers, Gregory M. Cooper

**Affiliations:** HudsonAlpha Institute for Biotechnology, Huntsville, AL, 35806, USA; Department of Medical Sciences, University of Torino, 10126 Torino, Italy; Molecular Biotechnology and Health Sciences Dept., Università degli Studi di Torino, via Quarello 15, 10135 Torino, Italy; Molecular Diagnostic Laboratory, Greenwood Genetic Center, Greenwood, SC, 29646, USA; University of Lausanne, Jules Gonin Eye Hospital, Fondation Asile des Aveugles, Lausanne, Switzerland; Department of Genetic Medicine and Development, University of Geneva, Switzerland; Obestetrics and Gynecology Department, Golestan University of Medical Sciences, Gorgan, Iran; Institute of Medical Genetics, University of Zurich, Schlieren 8952, Switzerland; Rare Diseases and Medical Genetics Unit, Bambino Gesù Children’s Hospital, IRCCS, Rome, Italy; Univ Brest, Inserm, EFS, UMR 1078, GGB, F-29200 Brest, France; GeneDx, LLC, Gaithersburg, MD, 20877, USA; Vanderbilt University Medical Center, Nashville, TN, 37232, USA; Department of Genetics, University of Alabama at Birmingham, Birmingham, AL, USA; Pediatrics and Medical Genetics, University of Colorado, Aurora CO, USA; Service de Médecine Génomique des Maladies de Système et d’Organe, Département Médico-Universitaire BioPhyGen, Hôpital Cochin, APHP, Université Paris Cité, Paris, France; Genetic Medicine, UCSF/Fresno, Fresno, CA, 93701, USA; Neuroscience Research Center, Faculty of Medicine, Golestan University of Medical Sciences, Gorgan, Iran; Nikagene Genetic Diagnostic Laboratory, Gorgan, Golestan, Iran; Department of Clinical Genetics, Erasmus MC University Medical Center, Rotterdam, The Netherlands; Boston Children’s, Boston, MA, USA; Children’s Hospital of Eastern Ontario Research Institute, Ottawa, Ontario, Canada; Service de Génétique Moléculaire et Génomique, CHU, Rennes, F-35033, France; Univ Rennes, CNRS, IGDR, UMR 6290, Rennes, F-35000, France; Children’s Mercy Kansas City, Center for Pediatric Genomic Medicine, Kansas City, KS, USA; Children’s Medical Center, Dallas TX, USA; Medical Genetics, Institute of Medical Genetics and Pathology, University Hospital Basel, Basel, Switzerland; Research Unit for Rare Diseases, 1st Faculty of Medicine, Charles University in Prague, Czech Republic; Department of Pediatrics and Inherited Metabolic Disorders, 1st Faculty of Medicine, Charles University in Prague, Czech Republic; Genetics and Rare Diseases Research Division, Ospedale Pediatrico Bambino Gesù, IRCCS, 00146, Rome, Italy; Medical Genetics Unit and Thalassemia Center, San Luigi University Hospital, University of Torino, Orbassano, Italy; Center for Integrative Genomics, University of Lausanne, Lausanne, Switzerland; Childrens Hospital of Philadelphia, Philadelphia PA, USA; Section of Genetics & Metabolism, Department of Pediatrics, University of Colorado Anschutz Medical Campus, Aurora, CO 80045; Division of Genetics and Metabolism, Children’s Health, Dallas, TX, USA; Pathology & Laboratory Medicine, Genomic Medicine, Neurological and Pediatrics Institutes, Cleveland Clinic, Cleveland, OH, USA; Service de Génétique Clinique, Centre de Référence Maladies Rares CLAD-Ouest, CHU Hôpital Sud, Rennes, France; Department of Pharmacology and Toxicology, College of Pharmacy, University of Utah, Salt Lake City, UT 84112, USA; University Children’s Hospital Zurich, University of Zurich, Zurich 8032, Switzerland; Service de Génétique Médicale et Biologie de la Reproduction, CHU de Brest, France; Centre de Référence Déficiences Intellectuelles de causes rares, Brest, France; Division of Genetics, Children’s Mercy Kansas City, Kansas City, MO; Genetics and Metabolism, Arkansas Children’s Hospital, Little Rock, AR, 72202, USA; Department of Pediatrics, Division of Genetics and Metabolism, University of Texas Southwestern Medical Center, Dallas TX, USA; Department of Endocrinology and Genetics, University Clinic for Children’s Diseases, Medical Faculty, University Sv. Kiril i Metodij, Skopje, Republic of Macedonia; Amsterdam University Medical Centers, Department of Clinical Genetics, Amsterdam, The Netherlands; Department of Human Genetics, Amsterdam University Medical Centers, VU University Amsterdam; Amsterdam Neuroscience, Amsterdam, The Netherlands

**Keywords:** *ZMYM3*, X-linked intellectual disability, neurodevelopmental disorder, transcriptional coregulators, chromatin modifiers

## Abstract

Neurodevelopmental disorders (NDDs) often result from highly penetrant variation in one of many genes, including genes not yet characterized. Using the MatchMaker Exchange, we assembled a cohort of 22 individuals with rare, protein-altering variation in the X-linked transcriptional coregulator gene *ZMYM3*. Most (n=19) individuals were males; 15 males had maternally-inherited alleles, three of the variants in males arose *de novo*, and one had unknown inheritance. Overlapping features included developmental delay, intellectual disability, behavioral abnormalities, and a specific facial gestalt in a subset of males. Variants in almost all individuals (n=21) are missense, two of which are recurrent. Three unrelated males were identified with inherited variation at R441, a site at which variation has been previously reported in NDD-affected males, and two individuals have *de novo* variation at R1294. All variants affect evolutionarily conserved sites, and most are predicted to damage protein structure or function. *ZMYM3* is relatively intolerant to variation in the general population, is highly expressed in the brain, and encodes a component of the KDM1A-RCOR1 chromatin-modifying complex. ChIP-seq experiments on one mutant, ZMYM3^R1274W^, indicate dramatically reduced genomic occupancy, supporting a hypomorphic effect. While we are unable to perform statistical evaluations to support a conclusive causative role for variation in *ZMYM3* in disease, the totality of the evidence, including the presence of recurrent variation, overlapping phenotypic features, protein-modeling data, evolutionary constraint, and experimentally-confirmed functional effects, strongly supports *ZMYM3* as a novel NDD gene.

## INTRODUCTION

Neurodevelopmental disorders (NDD) as a group affect 1-3% of children, but individual NDDs are typically rare and often result from highly penetrant genetic variation affecting one of many NDD-associated loci^1,2^. While exome/genome sequencing tests have provided molecular diagnoses for many individuals, the diagnostic yield of NDDs with these tests remains below 50%^3^. Various hypotheses exist to explain this diagnostic limitation, one of which is that many NDD-associated genes have yet to be identified. The wide availability of sequencing tests, coupled with data sharing, has allowed identification of many new NDD genes over the last few years^4^.

X-linked intellectual disability (XLID) is a subset of NDDs that accounts for a portion of the observed increased ratio of male to female probands with NDDs^5^. However, it can be difficult to separate pathogenic and benign variation on the X chromosome since many pathogenic XLID variants are inherited from unaffected heterozygous mothers^6,7^. For autosomal dominant ID genes, in contrast, pathogenic variation frequently arises *de novo*, and this fact can be used for both robust quantification of global disease enrichments and as a key piece of evidence for clinical interpretation of individual variants^8^. Despite this, data from large sequence databases such as gnomAD^9^ and TopMed/BRAVO (https://bravo.sph.umich.edu/freeze8/hg38/) do provide critical frequency data from control populations to support (or refute) new XLID associations^6^.

*ZMYM3* (MIM: 300061) lies on the X chromosome and encodes a member of a transcriptional corepressor complex that includes HDAC1, RCOR1, and KDM1A^10,11^. *ZMYM3* was originally identified as an XLID candidate gene in a female with a balanced X;13 translocation affecting the 5’ UTR of one isoform of *ZMYM3*^12^. The proband presented with ID, scoliosis, spotty abdominal hypopigmentation, slight facial asymmetry, clinodactyly, and history of a possible febrile seizure at age one year. Additionally, Philips, *et al*. reported a family with three NDD-affected brothers carrying a missense variant in ZMYM3 (R441W)^13^. The brothers displayed developmental delay, a sleeping disorder, microcephaly, genitourinary anomalies and facial dysmorphism.

Given the extremely low prevalence for any given Mendelian NDD, data sharing to facilitate cohort building is essential and has had a large impact on rare disease gene discovery over the last decade^14^. Here we describe a cohort of individuals with rare variants in *ZMYM3*, assembled from submissions to GeneMatcher^15^ and PhenomeCentral^16^. We provide strong evidence for an X-linked, *ZMYM3*-associated NDD based on phenotypic, computational, and experimental analysis of variants observed in 22 individuals.

## SUBJECTS AND METHODS

*ZMYM3* was submitted to GeneMatcher (https://genematcher.org/) by HudsonAlpha in 2018, and follow-up discussion of cases from either research studies or clinical sequencing was performed via email over the course of four years. Some matches originated from GeneMatcher^15^, while others originated from PhenomeCentral^16^. Over the course of the collaboration, some affected individuals were excluded from the cohort due to segregation of the variant of interest in unaffected male family members, including individuals harboring R688H, a variant that was initially identified as a VUS but later reclassified to likely benign after observation in an unaffected male relative. Additionally, one of the individuals with R688H variation did not have a phenotype that was similar to those described here; he had normocephaly, ID, autism, developmental regression and facial dysmorphism dissimilar to those described here.

Approval for human subject research was obtained from all local ethics review boards, and informed consent for publication (including photos, where applicable) was obtained at individual sites. Exome sequencing (ES), genome sequencing (GS), or panel testing was performed as described in Supplemental Materials and Methods.

For protein modeling, the wild-type 3D protein structure was downloaded from AlphaFoldDB (https://alphafold.ebi.ac.uk/)^17^, which was included with the reference from UniProt (Accession number: Q14202). When not possible online, structures were visualized, colored, and the sequence was mutated with Chimera version 1.15, rotamer builder tool^18^. Structure superposition was obtained in Chimera with the tool Matchmaker. Structure refinement was performed with the Chimera tool Dock Prep with standard settings, as previously described^19^. Depiction of molecular surfaces was defined as VdW surface and colored according to the electrostatic potential. For additional analyses, see Supplemental Materials and Methods.

For ChIP-seq experiments, we edited the genomic DNA at the *ZMYM3* endogenous locus in HepG2 cells to introduce the variant (the “variant” experiment) or to reintroduce the reference sequence (the “control” experiment), simultaneously with a 3X FLAG tag, 2A self-cleaving peptide, and neomycin resistance gene, using a modified version of the previously published CRISPR epitope tagging ChIP-seq (CETCh-seq) protocol^20^. We nucleofected cells and selected for correctly edited cells using neomycin, confirmed edits by PCR and Sanger sequencing of genomic DNA, and performed ChIP-seq as previously described^21^ with duplicate experiments for each condition (see Supplemental Materials and Methods). We performed peak calling using SPP^22^ and Irreproducible Discovery Rate (IDR)^23^, using ENCODE-standardized pipelines for analysis and quality-control^24^. We performed additional differential binding analyses using the R package csaw v1.28.0^25^.

As an additional control, we used the standard CETCh-seq ZMYM3 experiment in HepG2 available on the ENCODE portal (ENCSR505DVB), with these data processed to match (i.e., downsampled to 20M reads) the other CETCh-seq experiments described here. See Supplemental Materials and Methods for additional details.

## RESULTS

### *ZMYM3* Variants

Through a collaboration facilitated by the MatchMaker Exchange^14^, we identified 17 unique variants in *ZMYM3* in 22 affected individuals from 20 unrelated families (Figure 1, Table 1). All observed variants had high CADD scores (average 24.2, range 19-32, Table S1), indicating that they rank among the most deleterious SNVs in the human reference assembly, similar to most known highly penetrant NDD-associated variants^26^.

**Table 1.**
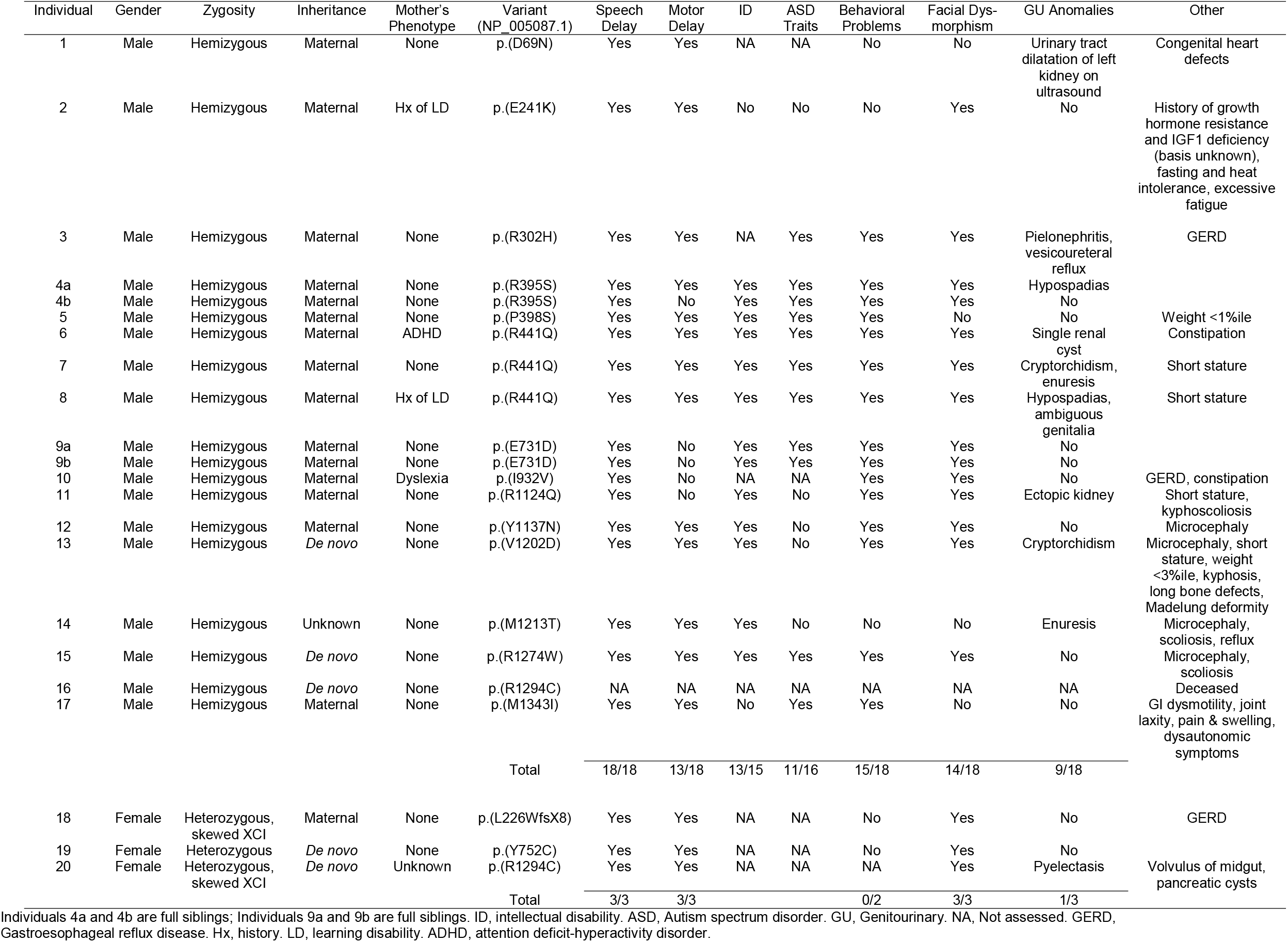
Individual variant and phenotypic data for the entire cohort.

**Figure 1.**
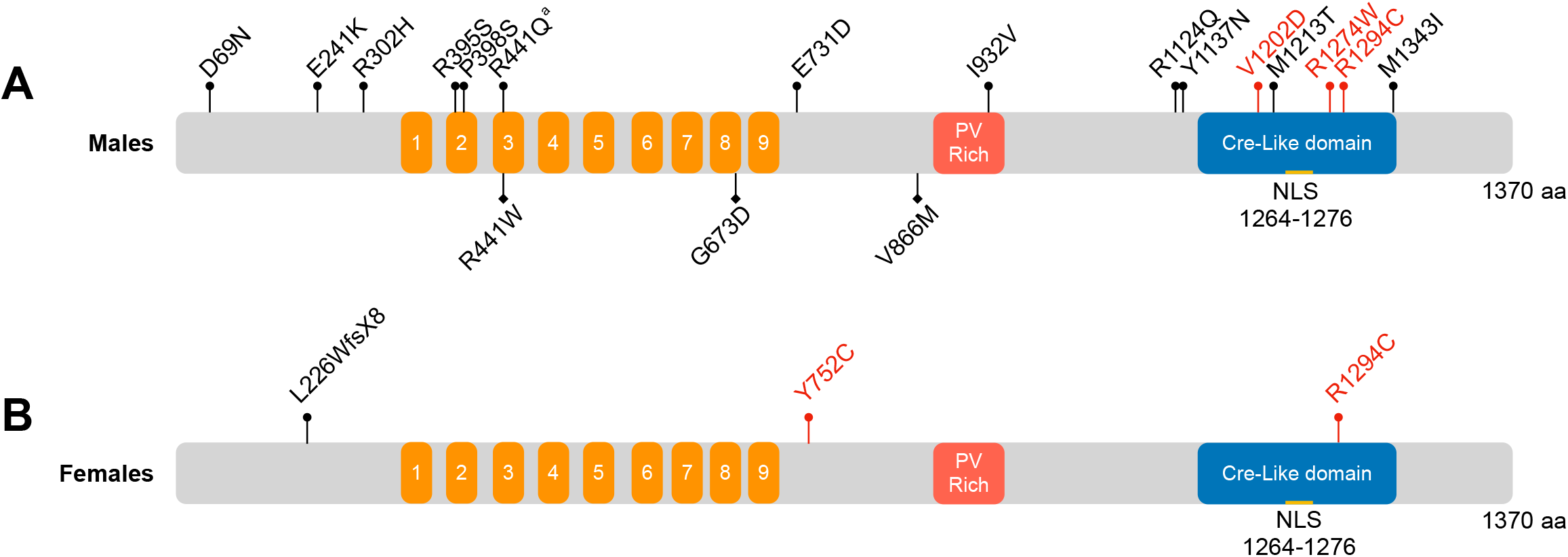
Observed variation along the length of the ZMYM3 protein. The 1370 aa ZMYM3 protein (Q14202, NP_005087.1) is annotated with MYM-type zinc fingers (1-9, orange), a Proline-Valine Rich region (PV Rich, red), and Cre-like domain (blue) as described by UniProt. **A**. Hemizygous variants observed in males in this study are noted above the protein model, with *de novo* variants in red. (a) Note that R441Q was observed in three unrelated males. Hemizygous variants that were previously reported in males are shown below the protein (from PMID: 32891193, PMID: 24721225). **B**. Maternally-inherited (black) or *de novo* (red) heterozygous variants observed in females in this study are noted above the protein model.

Nineteen of these 22 individuals are males that harbor hemizygous missense variants, including two sets of affected brothers. For the majority of males (n=15), variants were inherited from heterozygous carrier mothers. In three males, the *ZMYM3* variant arose *de novo*, while inheritance could not be defined for one. All variants were rare, with three or fewer total alleles and no hemizygous males or homozygous females in gnomAD^9^ or TopMed/Bravo (https://bravo.sph.umich.edu/freeze8/hg38/)(Table S1).

In addition, we identified three heterozygous *ZMYM3* variants in three unrelated, affected females (Figure 1, Table 1). All three variants are absent from population databases. Two of these variants arose *de novo*, while one was inherited from an apparently unaffected mother. In two of the three females, X-inactivation testing targeting either the *AR* locus^27^ or the *RP2* locus^28^ was performed, and in both, skewed X-inactivation was observed. In the case of the maternally-inherited L226WfsX8 variant, >94% skewing was observed in both the proband and her unaffected, heterozygous mother at the *RP2* locus. Both mother and daughter were heterozygous for two *RP2* alleles (366/362), and in both, the 366 allele was inactivated (see Supplemental Note: Case Reports for additional details). In Individual 20, a female carrying a *de novo* R1294C variant, 97% skewing at the *AR* locus was observed. Skewing of the precise *ZMYM3* alleles was not tested in these individuals.

### Phenotypic Characterization

Of the nineteen identified males, one died at 26 weeks gestational age with a *de novo* variant in *ZMYM3* (R1294C) and a very severe phenotype (Supplemental Note: Case Reports). For this reason, we did not include this male in further phenotypic comparisons. Of the remaining 18 affected males, all were reported to have developmental delay (18/18), with speech delay (18/18) being more prominent than motor delay (13/18)(Table 1, Supplemental Note: Case Reports). Of those who could be assessed, 13/15 showed intellectual disability, and most were diagnosed with autism or were reported to have autistic traits (11/16). The majority of males had behavioral concerns at some point in development (15/18). Almost all affected males were also reported to have at least mild facial dysmorphism (14/18), some of which were highly similar to the individuals reported in Philips, *et al*.^13^(Figure 2). Similarities include thick eyebrows, deeply set eyes, long palpebral fissures, protruding ears, and a high anterior hairline. Other variable features include genitourinary anomalies (9 individuals), short stature (4), microcephaly (4), scoliosis/kyphosis (4), and functional gastrointestinal problems (5) (Table 1). See Supplemental Note: Case Reports for additional clinical features for each case.

**Figure 2.** Facial features of a subset of individuals with *ZMYM3* variation. Individual ID and protein effect are noted for each. Note deep-set eyes, long palpebral fissures, large/prominent/cupped ears, and tall forehead.

While most variants in males were inherited from unaffected heterozygous carrier mothers (9/13 mothers), the remaining four heterozygous mothers were reported to have a history of learning disabilities, attention deficit-hyperactivity disorder (ADHD), or dyslexia (Table 1, Supplemental Note: Case Reports, Figure S1).

Among the affected females, all three displayed developmental delay and some facial dysmorphism, but many of their additional features were variable and do not lead to a clear syndromic picture (Table 1, Figure 2, Supplemental Note: Case Reports).

### Protein Modeling

*ZMYM3* encodes a DNA-binding transcriptional coregulator with multiple protein isoforms, the longest of which is 1370 amino acids (Q14202, NP_005087.1). This isoform has nine MYM-type zinc fingers, a proline-rich region, and a C-terminal Cre-Like domain (Figure 1). As most of the observed variants are missense (16/17 unique variants), we performed computational modeling to assess the potential effects of these changes. Homology-based protein modeling using AlphaFold^17^ indicates that 13 of the 16 missense variants lie in ordered regions, and the majority have intermediate to high predicted Local Distance Difference Test (pLDDT) scores^29^, indicating that there is a moderate to high degree of confidence in further computational predictions (Figure 3, Figure S2, Table S2).

**Figure 3.**
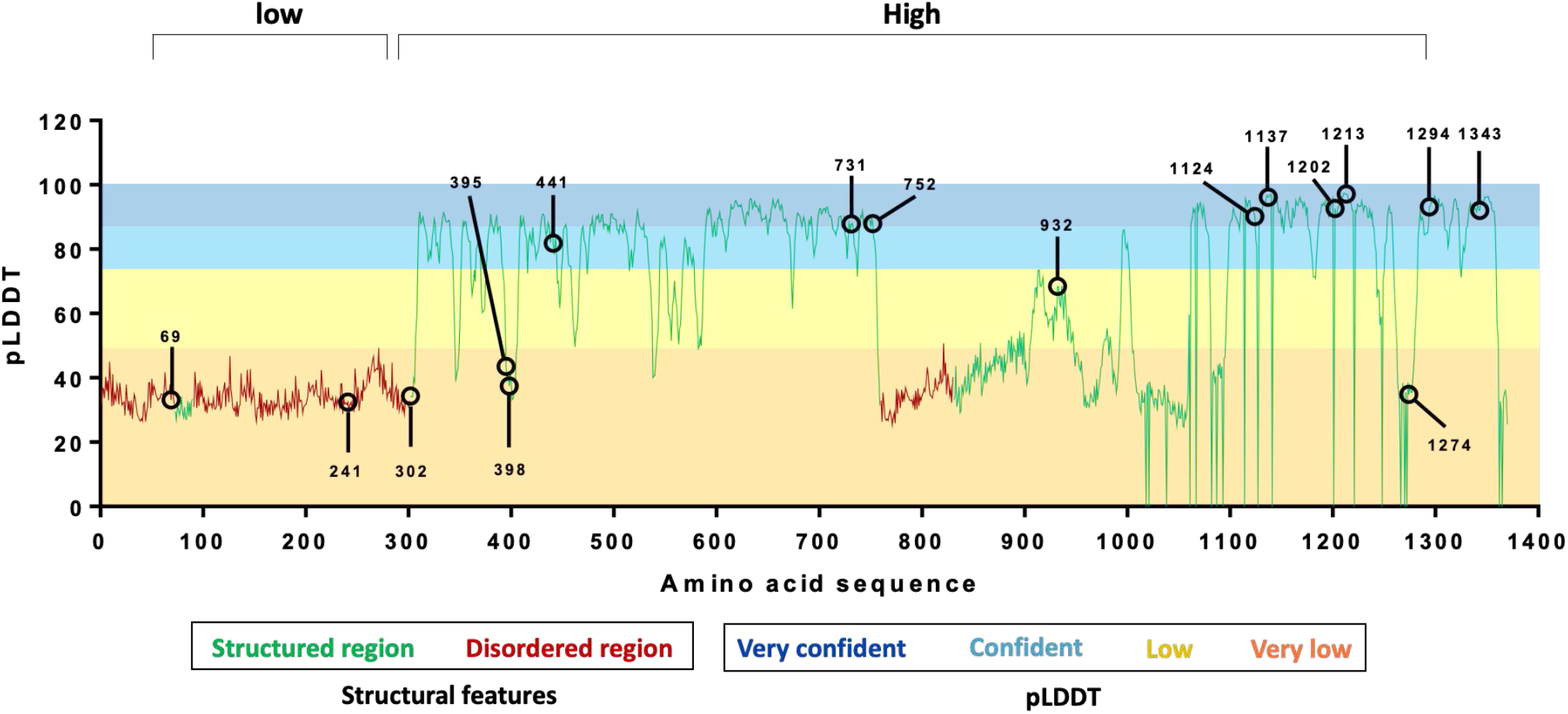
Missense variants in ZMYM3 mainly lie in ordered regions. Three disordered regions (red line) were identified (aa 1-72, 90-301, and 759-830), while the remainder of the protein is predicted to be structured (green line). AlphaFold produces a per-residue confidence score (predicted Local Distance Difference Test, pLDDT) between 0 and 100, which is plotted along the length of the ZMYM3 protein. Horizontal bars and shading indicate confidence ranges for pLDDT scores. Missense variants observed here are noted on the graph, and while residues 69, 241, and 302 lie in disordered regions, the remainder of residues lie in structured regions.

We assessed flexibility, stability, solvent exposure, and deformation energy of the mutant protein models (Figures S3-S6). A general trend towards protein destabilization (negative folding energy differential) was observed for several mutants, while R1274W was predicted to be stabilizing (Figure S3). We observed patterns somewhat consistent with solvent exposure across the 16 unique missense variants (Table S2). Six of the seven variants leading to the highest destabilization (R441Q, E731D, Y752C, R1124Q, Y1137N, M1213T) represent buried residues in high confidence regions of the protein. While disruption of each of these rigid residues is predicted to be destabilizing, some are due to likely increased flexibility (E731D, Y752C, Y1137N) while others are predicted to be more rigid (R441Q, R1124Q, M1213T). This result is consistent with the observation that substitutions of amino acids within the protein core are often associated with folding destabilization.

Conversely, the remaining 10 variants are exposed residues; six of these lie in low confidence regions or have very low pLDDT values (D69N, E241K, R302H, R395S, P398S, R1274W). These wild type residues are predicted to be flexible, and in most cases the observed mutation is predicted to lead to a more rigid structure. The remaining four residues represent rigid residues that have varying predicted deleterious effects. More detailed surface analyses indicated that several variants result in significant changes of polarity, charge, and hydrophobicity (Table S2, Figure S6). In particular, R1274W is predicted to have major effects, resulting in stabilization of an exposed residue through the substitution of a polar, charged and flexible arginine with a neutral, aromatic and hydrophobic tryptophan moiety (Figure S6).

In addition to structural analysis, we submitted the sequences to the Eukaryotic Linear Motif (ELM)^30^. This resource predicts short amino acid motifs either bound by other proteins, or sites of post-translational modifications (phosphorylation, cleavage sites, ubiquitination, etc.). Intersecting this information with the position of our mutations suggests that several of the variants alter motifs (Table S2, Table S3) and that modifications of residues R302, V1202, M1213, R1274, and M1343 are predicted to possibly disrupt multiple interactions.

### Genome-wide occupancy of selected ZMYM3 variant transcription factors

A key role of ZMYM3 is to function as a component of the KDM1A/RCOR1 chromatin-modifying complex that regulates gene expression^10^. Therefore, we sought to measure the impact of variation on ZMYM3 activities genome-wide. Given the time and expense of these experiments, we chose three variants for testing: R441W, a previously reported variant^13^ that affects a residue where we have seen recurrent variation (R441Q); R1274W, a *de novo* variant within the Cre-like domain that was found in an individual with notable facial similarities to those individuals with R441 variation; and R688H, which early in our collaboration appeared to be a recurrent variant seen in two affected individuals. Subsequently, segregation studies in one family indicated that the R688H variant was present in an unaffected maternal uncle, suggesting that it is likely benign.

For each of these, we introduced the variant into the *ZMYM3* gene in the genomic DNA of cultured HepG2 cells using a modified version of the CRISPR epitope tagging ChIP-seq (CETCh-seq) protocol^20^; CETCh-seq uses an active Cas9 double-stranded DNA cleavage followed by homology-directed repair with an introduced donor template. In these experiments, we simultaneously introduced a “super-exon” consisting of all exons of *ZMYM3* downstream (relative to coding direction) of the exon in which the variant resides, along with a FLAG epitope tag and selectable resistance gene. These modifications result in cells that express the ZMYM3 protein with the variant residue and a carboxyl-terminus FLAG tag for immunoprecipitation, as well as a neomycin resistance gene product for selection (cells that survive selection contain an in-frame insertion of the neomycin resistance gene that is driven by the endogenous *ZMYM3* promoter and indicates the transcription of in-frame *ZMYM3* with the FLAG tag). As a control for each super-exon edit, we performed the same protocol on cells but reintroduced the reference sequence in the super-exon along with the FLAG tag and resistance gene. In both the control and variant experiments, an additional single base substitution was introduced, located in the Protospacer Adjacent Motif (PAM) for the single guide RNA (sgRNA) used in the CRISPR editing, in order to abolish Cas9 cleavage after cells are correctly edited. The PAM mutation was either synonymous (for the ZMYM3^R688H^ and ZMYM3^R441W^ constructs) or intronic (for the ZMYM3^R1274W^ construct). The genomic DNA modifications were confirmed by PCR amplification and Sanger sequencing. The key advantage of this approach is that the control and variant ZMYM3 proteins are produced from the endogenous genomic loci, each modified by the same super-exon, and that the antibody used (along with other experimental and analytical steps) is the same; the only difference between the variant and control experiments is the presence of the missense variant of interest. For both R688H and R1274W, we successfully obtained correctly edited cells; however, for R441W, we were unable to obtain edited cells. For both R688H and R1274W, we performed chromatin immunoprecipitation followed by high-throughput sequencing (ChIP-seq) and peak calling as previously described^21,31^, downsampling all replicate experiments to 20 million reads to control for total read-depth (Table S4), and compared the genome-wide binding profiles of the control and variant experiments. As an additional control, we used a standard CETCh-seq experiment on ZMYM3 (ZMYM3^CETCh^) in HepG2 cells to establish baseline genome-wide occupancy of the wild-type ZMYM3 protein without a super-exon or missense variant (ENCODE dataset ENCSR505DVB).

When comparing ZMYM3^R1274W^-control (i.e. super-exon with reference sequence at the variant position) to ZMYM3^R1274W^-variant, we observed a large difference in the number of peaks called between the experiments, with the control experiment yielding 16,214 peaks and the variant only 3,699 peaks (Table S4). Among the variant peaks, 2,508 (67.8%) were also called in the control experiment. We know from extensive previous ChIP-seq analyses that many loci exhibit read-depth levels near (above or below) peak-calling thresholds, resulting in situations where experiments are more similar than they appear when only considering peak-call overlaps. Thus, we performed additional, more quantitative comparisons, including read-depth correlations and overlaps of peak-calls using both standard and relaxed thresholds; these analyses consistently show a large, global reduction of occupancy for R1274W variant experiments (see Supplemental Materials and Methods). The large disparity between the control and variant ZMYM3^R1274W^ experiments was also observed in a differential analysis using the R package *csaw*^25^. Rather than relying on peak calls, *csaw* performs a sliding window analysis to detect regions that show differing levels of read-depth between the experiments. Using the two replicates of ZMYM3^R1274W^-control and the two replicates of ZMYM3^R1274W^-variant, *csaw* identified 25,845 genomic regions with sufficient reads for analysis. Of these regions, 13,225 showed differential associations between control and variant at FDR < 0.05. All but 19 (99.9%) of these sites had higher read counts in the control than in the variant. We also intersected the 25,845 *csaw* regions with the union of peak calls between control and variant experiments (n=17,405), resulting in 11,259 genomic regions; of these, 4,625 were not differential at FDR < 0.05, while 6,631 were significantly higher in control than in variant, and only three were significantly higher in variant than in control (Figure 4A). We performed the *csaw*/peak overlap analyses between ZMYM3^CETCh^ and the R1274W variant and control experiments and observed that ZMYM3^CETCh^ and ZMYM3^R1274W^-control had very similar genomic association patterns, while ZMYM3^R1274W^-variant comprised only a small minority of ZMYM3^CETCh^ sites (Figure S7).

**Figure 4.**
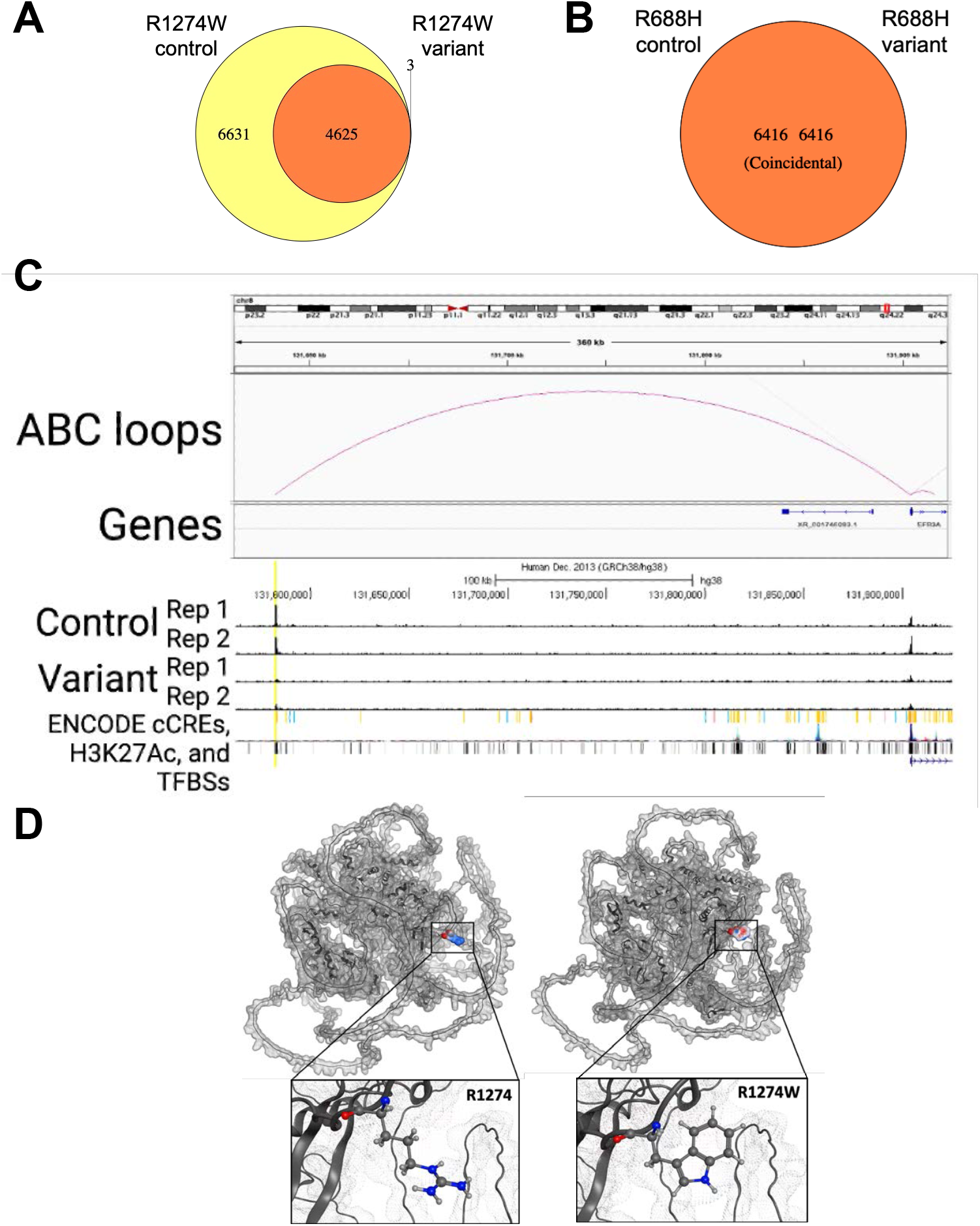
R1274W is a hypomorphic allele, while R688H has similar genome occupancy to that of wild type. **A, B**. Genomic regions called by csaw between experiments, then overlapped with IDR 0.05 peaks called in either experiment. Yellow color indicates regions determined by csaw to have significantly higher differential binding (at FDR < 0.05) in control, orange indicates regions with no differential binding, and red indicates regions with higher differential binding in variant. For R1274W (**A**), there are 6,631 regions with higher binding in control, 4,625 regions with no differential binding, and 3 regions with higher binding in variant. For R688H (**B**), all 6,416 regions had no differential binding. **C**. Genome browser track for ZMYM3-R1274W-variant ChiP-seq experiments. Human genome (hg38) chr8:131,561,953-131,925,404 is displayed. Top track is Activity-by-Contact (“ABC loops”) showing predicted interaction between enhancer element on left and TSS for the gene *EFR3A* on right. “Genes” track is RefSeq gene model. “Control Rep 1” and “Control Rep 2” are aligned bam reads from ZMYM3-R1274W-control experiments, “Variant Rep 1” and “Variant Rep 2” are aligned bam reads from ZMYM3-R1274W-variant experiments. All bam files are downsampled to an equal number of reads in each replicate, and all four replicate tracks are scaled from 0-60 vertically. “ENCODE” tracks are shown below the ChIP-seq tracks: “cCREs” represent candidate cis-regulatory elements colored by ENCODE standards, “H3K27Ac” is layered H3K27Ac signal from seven ENCODE cell lines, and “TFBSs” are ENCODE TF clusters (340 factors, 129 cell types). The putative enhancer element identified as the most significant loss of binding in the variant experiment is highlighted in yellow, showing the ENCODE distal enhancer cCRE call, the ABC loop to the TSS of *EFR3A*, and the difference in binding with the two control replicates showing strong signal and the two variant replicates showing substantially less binding. **D**. Protein modeling of R1274 and R1274W.

We similarly analyzed the ZMYM3^R688H^-control and ZMYM3^R688H^-variant experiments. Global correlation analyses indicated a high degree of similarity of genomic occupancy between the control and variant experiments (Supplemental Materials and Methods). Analysis of *csaw* regions intersected with peak calls gave 6,416 genomic sites, none of which were significantly different (FDR < 0.05) between control and variant (Figure 4B). Pairwise Pearson correlation coefficients of read counts of each of the two replicates of control and variant ranged from 0.71 to 0.84, indicating a high degree of overall similarity (Figure S8). However, both control and variant R688H experiments yielded less binding and fewer peaks than both ZMYM3^R1274W^-control and ZMYM3^CETCh^ experiments (Figure S7). We hypothesize that the super-exon edit itself is affecting the ZMYM3^R688H^ function. Importantly, we find no substantial difference between control and variant function, supporting the hypothesis that R688H is a likely benign variant.

## DISCUSSION

Here we describe 22 NDD-affected individuals with protein-altering variation in *ZMYM3*. While four of these variants arose *de novo*, most were present in males in a hemizygous state and inherited from unaffected or mildly affected heterozygous mothers. All variants presented here are rare and predicted to be deleterious. Many of the variants are predicted to interfere with protein structure or function. *ZMYM3* is relatively intolerant to both missense variation (gnomAD missense Z=4.31) and loss-of-function variation (RVIS=8.46^32^, pLOEUF=0.11^9^), further supporting the potential for the variants observed here to have phenotypic effects, and suggesting that the gene is likely dosage sensitive. Using ChIP-seq, we have also provided functional analyses showing that one deleterious variant, R1274W, acts as a hypomorphic allele, with reduced genome occupancy compared to its control and to wild type, and that one likely benign allele, R688H, has genome occupancy similar to its control experiment.

Among the variants in our cohort, there are two sets of alleles affecting the same codon. At R441, a residue that lies within a zinc finger domain that functions in DNA binding, we found substitutions (R441Q) in three unrelated males. Three additional affected males with a distinct allele at this same residue (R441W) in one family have been previously reported^13^. Overlapping phenotypic features of these six individuals include developmental delay (mainly speech), nocturnal enuresis, and microcephaly. In addition, the facial features in these R441 individuals are quite similar. The other recurrent variant that we observed here is R1294C, observed as *de novo* in a deceased male and in a female with 97% skewed X-inactivation. R1294C has also been submitted to ClinVar^33^ as a VUS (SCV000297052.2) by a different group than those that identified R1294C variation for this study. We thus believe the ClinVar submission represents a third, independent R1294C case, although we are unable to confirm this (see Supplemental Materials and Methods).

The biological role of *ZMYM3* is also supportive of disease relevance. ZMYM3 is part of a transcriptional corepressor complex that includes HDAC1, RCOR1 and KDM1A^10,11^. Additional interactors in this complex can include ZMYM2 and REST. Variation in two of these five genes has been robustly associated with neurodevelopmental disorders^34,35^. Additionally, ZMYM3 has since been shown to physically interact with RNASEH2A; variation in *RNASEH2A* (MIM:606034) has been associated with Aicairdi-Gouteres Syndrome 4 (AGS4, MIM:610333). Specifically, a cluster of pathogenic variants found in AGS4 patients have been shown to disrupt binding of RNAseH to ZMYM3^11^. Residues within the PV-rich domain of ZMYM3 (specifically 862-943) have been shown to be necessary for this interaction. I932V, observed in our cohort, lies in this region and may disrupt this interaction.

Recently, Connaughton *et al*. demonstrated a connection between loss-of-function variation in *ZMYM2* (MIM:602221), a paralog of *ZMYM3* with 44% protein-identity, to congenital anomalies of the kidney and urinary tract, with extra-renal features or NDD findings (MIM:619522)^34^. This same publication also reported two male probands who had hemizygous variants of uncertain significance in *ZMYM3*, resulting in G673D and V866M (Figure 1). Phenotypic overlap of individuals with variation in *ZMYM2* and *ZMYM3* presented here include developmental delay, microcephaly, and ID. Some similarity of facial features is also shared with the *ZMYM2* cohort, including one proband with protuberant ears. In addition to ZMYM2 and ZMYM3, the ZMYM-family of proteins includes two additional members, ZMYM4, and QRICH1. Variation in *QRICH1* (MIM:617387) has been associated with Ververi-Brady syndrome (MIM:617387), which has features including developmental delay, intellectual disability, non-specific facial dysmorphism, and hypotonia among others^36^.

Variants observed in this cohort lie across the length of the protein, and modeling data suggest that while several may affect protein structure, several also likely affect protein-interactions, which are key in the biological function of ZMYM3. ChIP-seq data for ZMYM3^R1274W^ indicate a large reduction in genome-wide occupancy, even though the variant is not within any direct DNA-binding domains. Leung *et al*. have previously shown that this specific residue is necessary for interaction with RAP80, a ubiquitin-binding protein that plays a role in the DNA damage response^37^. While the observed widespread reduction in genomic occupancy indicates a global hypomorphic effect, individual binding event differences may be of particular interest. For example, one of the most significant differential binding events, as determined by *csaw*, occurs at a regulatory element on chromosome 8 (Figure 4C); this region is annotated as a distal enhancer by the ENCODE Consortium^38^, and, according to Activity-by-Contact (ABC) analysis^39^, this region connects to and is likely a regulatory element for the gene *EFR3A*. Pathogenic variants in *EFR3A* have been associated with autism spectrum disorders^40^, with phenotypes that overlap those described here.

A key limitation of this study is the X-linked nature of *ZMYM3*, and the fact that most of the variants observed here are inherited, which makes the statistical evaluation of pathogenicity difficult. We cannot, for example, use *de novo* variant enrichment testing, a powerful means of inferring pathogenicity for dominant NDDs^41^. Traditional association or burden testing also cannot be done given the absence of systematically ascertained and matched cases and controls. However, this is a common limitation of establishing X-linked recessive disease in males. Additionally, while many pedigrees suggest that testing in other family members may be informative for each individual variant’s interpretation (Figure S1), we were unable to test all siblings and relatives. This additional information may be particularly useful for flagging any potential benign variants within these families (i.e., those present in unaffected male relatives, such as was observed for R688H). X-chromosome inactivation studies in carrier females may also be informative.

Despite the above limitations, the totality of the evidence presented here is strong. The presence of recurrent variation, overlapping phenotypic features, protein-modeling data, evolutionary constraint, and experimentally-confirmed functional effects strongly support *ZMYM3* as a novel NDD gene. While additional analyses are necessary to ultimately confirm these findings and adjudicate the pathogenicity of each individual variant, we provide substantial evidence that *ZMYM3* is likely to be a novel NDD-associated gene.

## Data Availability

The published article includes all variant information pertinent to this study. ChIP-seq data will be made available via the NCBI Gene Expression Omnibus (GEO, https://www.ncbi.nlm.nih.gov/geo/).

## Acknowledgements

We sincerely thank all the families who participated in this study.

SMH, AC, ACEH, MD and GMC were supported by a grant from the Alabama Genomic Health Initiative (an Alabama-State earmarked project F170303004) through the University of Alabama in Birmingham. LN was supported by the project National Institute for Neurological Research (Programme EXCELES, ID Project No. LX22NPO5107), Funded by the European Union, Next Generation EU and by institutional program UNCE/MED/007 of Charles University in Prague. SK was supported by grant NV19-07-00136 from the Ministry of Health of the Czech Republic. LN and SK thank the National Center for Medical Genomics (LM2018132) for WES analyses. Sequencing and analysis of one individual in this study was made possible by the generous gifts to Children’s Mercy Research Institute and Genomic Answers for Kids program at Children’s Mercy Kansas City. This work was also supported by the Italian Ministry of Health (Ricerca 5×1000, RCR-2020-23670068_001, and RCR-2021-23671215 to MT), and Italian Ministry of Research (FOE 2019, to MT), and PRIN2020 (code 20203P8C3X, to AB). Reanalysis of exome sequencing for individual 14 was performed on a research basis by the Care4Rare Canada Consortium. This work was supported by grants from the Swiss National Science Foundation (31003A_182632 to AR) and the Blackswan Foundation (to AR), and the ChildCare Foundation to SEA. This work was also funded through the CRT Foundation (Progam “Erogazioni Ordinarie” 2019) and the Italian Ministry of University and Research (Assegni, Tornata 2022, Bando: BMSS.2022.06/XXIV), to MRS, GC and GE. MM was supported by RVO VFN 64165, Czech Ministry of Health. This work was also supported by a Swiss National Science Foundation grant 320030_179547 to AR. CN and JD were supported by The Genesis Foundation for Children.

