## SupplementalFigures for "Deleterious, protein-altering variants in the X-linked transcriptional coregulator *ZMYM3* in 22 individuals with a neurodevelopmental delay phenotype"

**Figure S1.** Pedigrees for each family in the study. Variants are shown above the pedigree, with *de novo* variation in red. Affected individuals presented here are indicated with arrows and labeled with IDs matching the text (1-20). Individuals affected with an NDD are shown in black, while mildly-affected individuals, those with a single feature of an NDD, or a history of such are shown in gray. Presence of the variant is indicated with +, while absence of the variant (if tested) is indicated by a -. Numbers in unaffected individuals' shapes indicate the number of unaffected siblings (or half-siblings) when there are more than two.

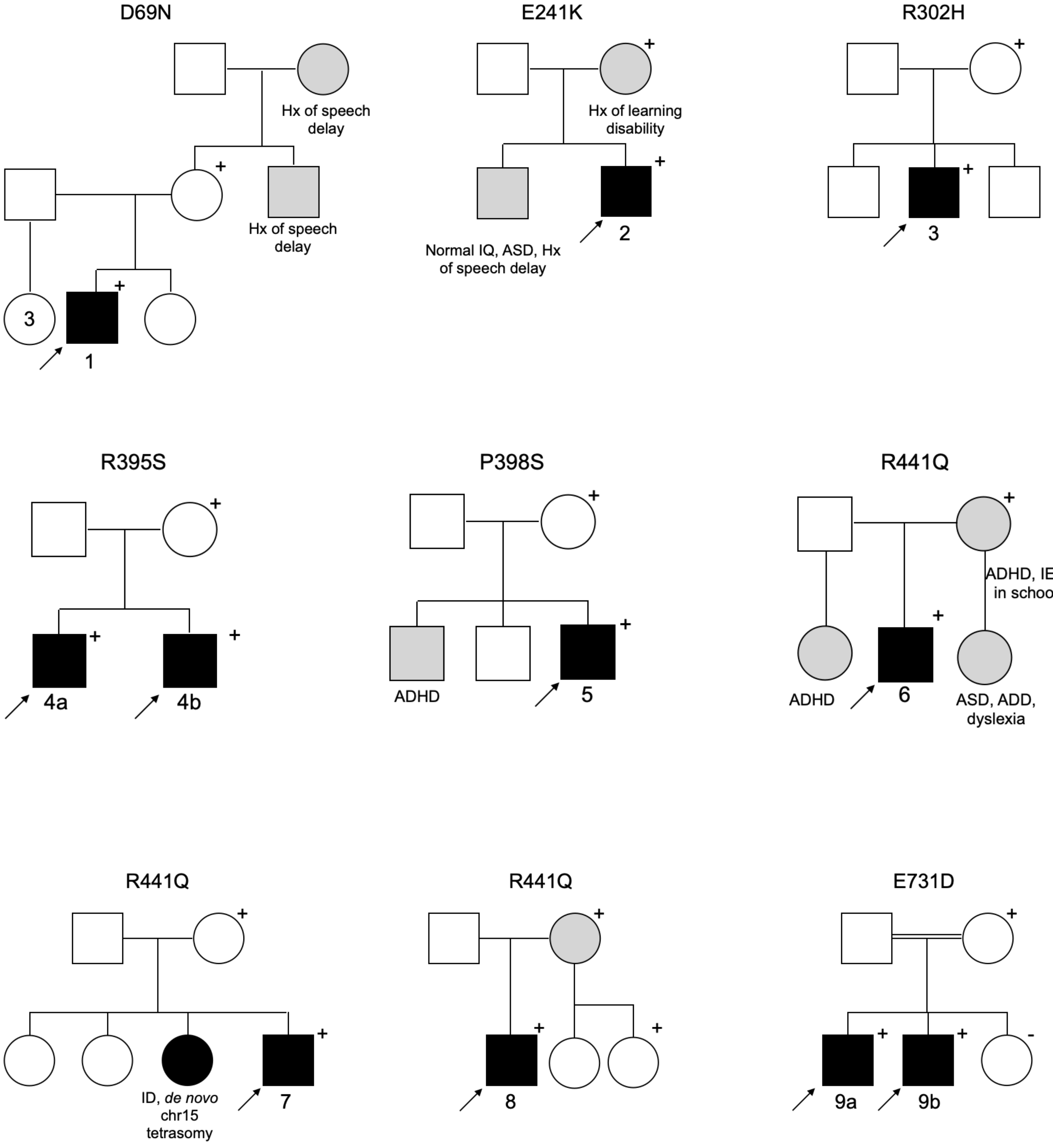

I932V

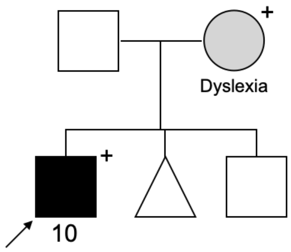


R1124Q

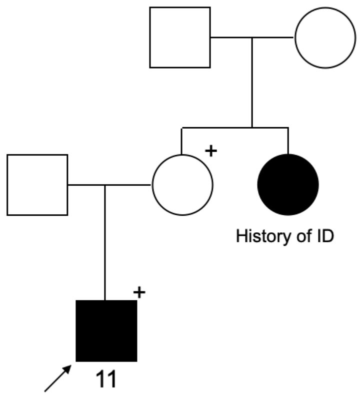


Y1137N

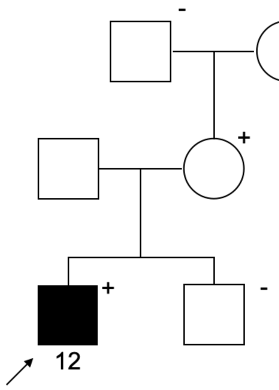


V1202D

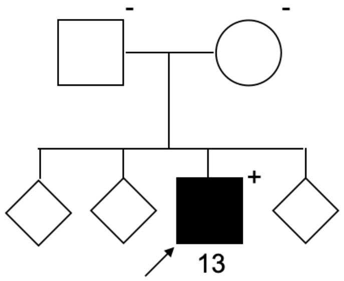


M1213T

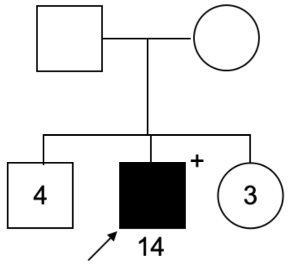


R1274W

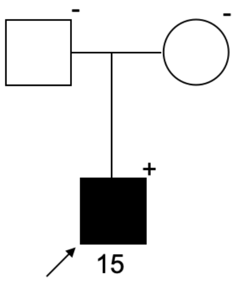


R1294C

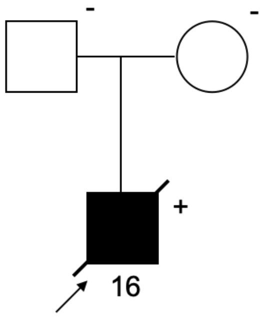


M1343I

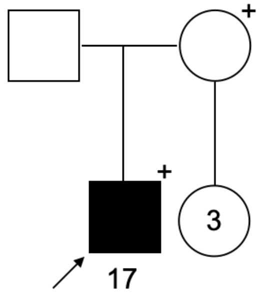

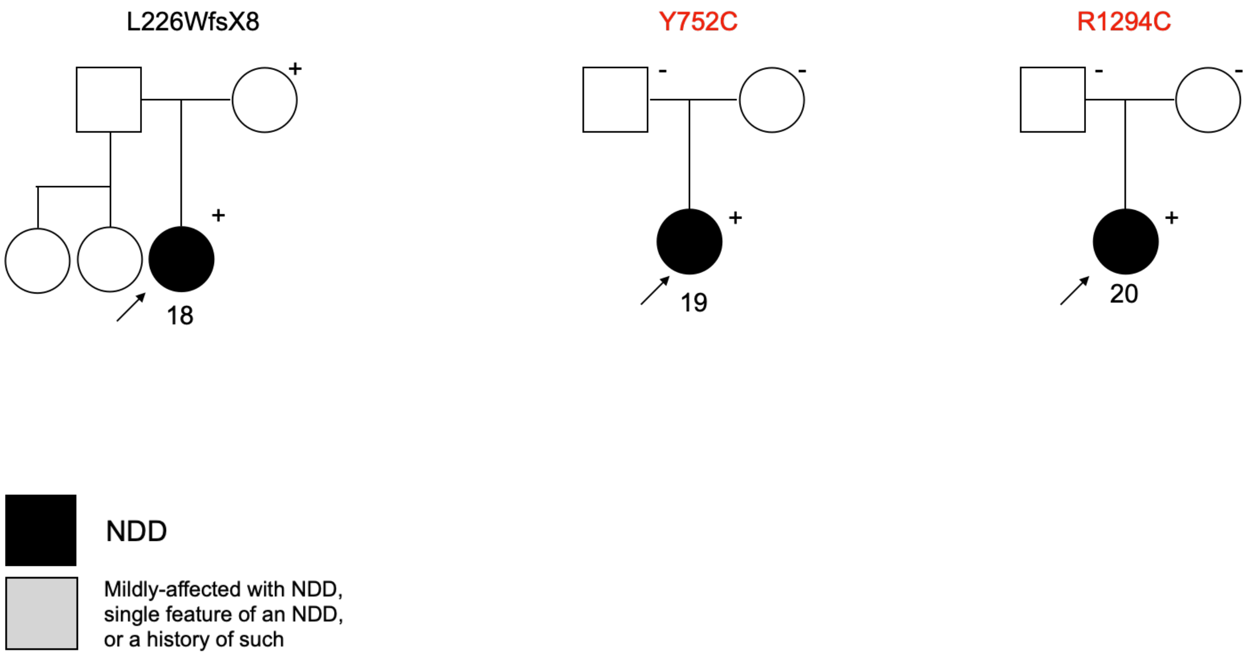
L226WfsX8
+
+
18
Y752C
-
-
+
19
R1294C
-
-
+
20
NDD
Mildly-affected with NDD,
single feature of an NDD,
or a history of such

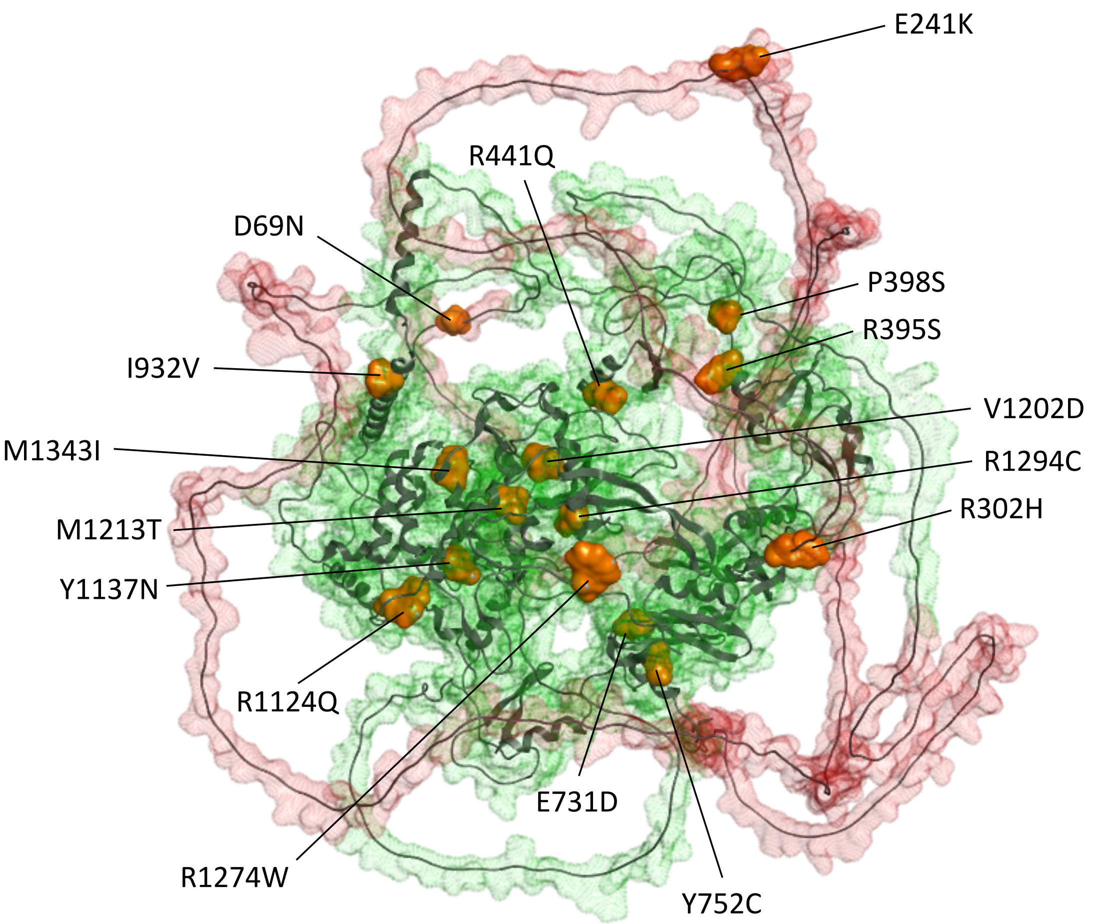


**Figure S2. Missense variants in ZMYM3 mainly lie in ordered regions.** Visual representation of the variant residues on a 3D model of ZMYM3. Disordered regions are in red, while structured regions are shown in green. Individual variants are shown in orange.

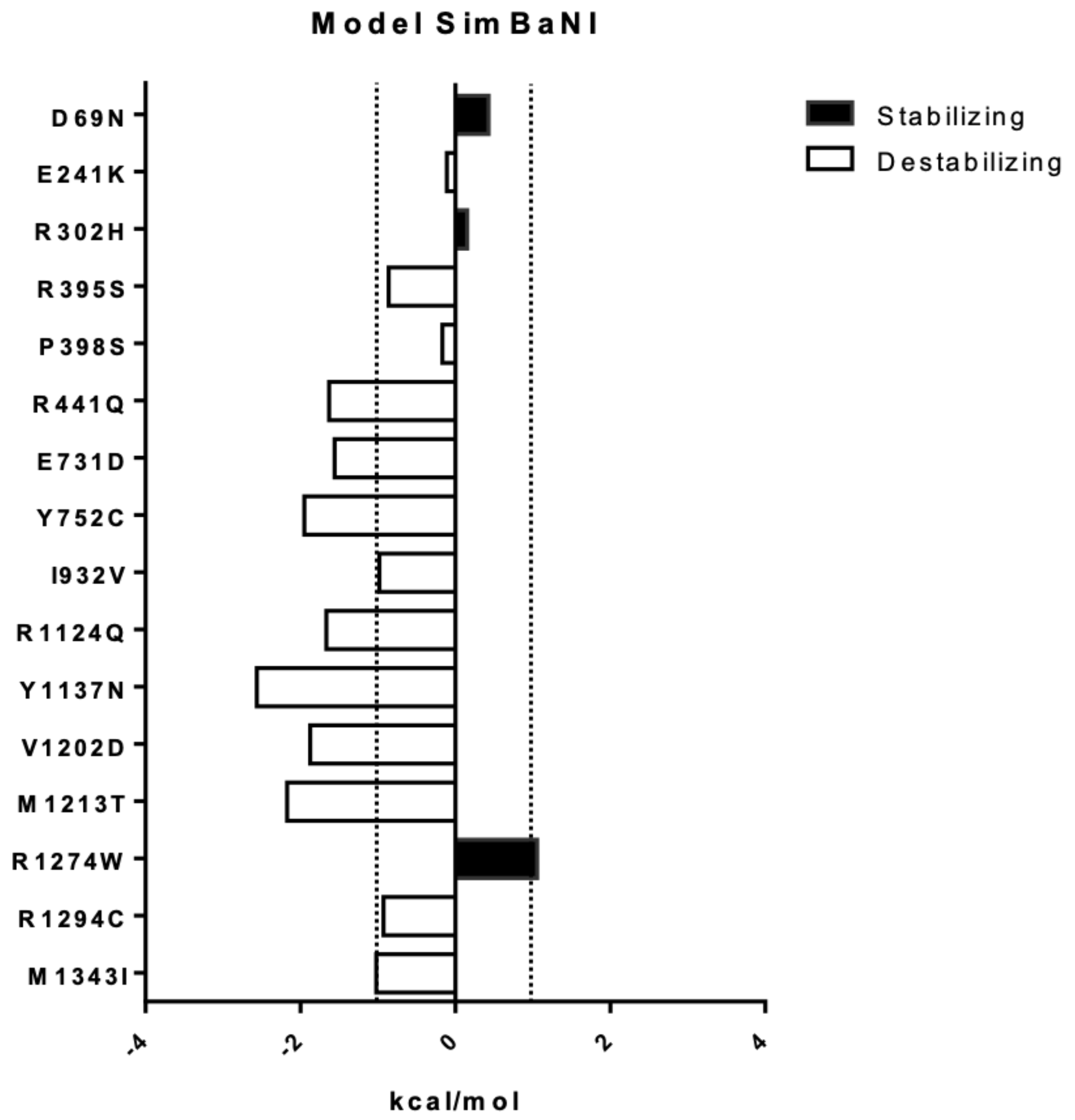


**Figure S3.** Analysis of the free energy variation upon amino acid substitution suggests a destabilization for the majority of mutants with a major impact of mutants R441Q, E731D, Y752C, R1124Q, Y1137N, V1202D, and M1213T. Conversely, R1274W is predicted to stabilize the protein structure. Vertical dotted line indicates threshold of +/- 1 kcal/mol.

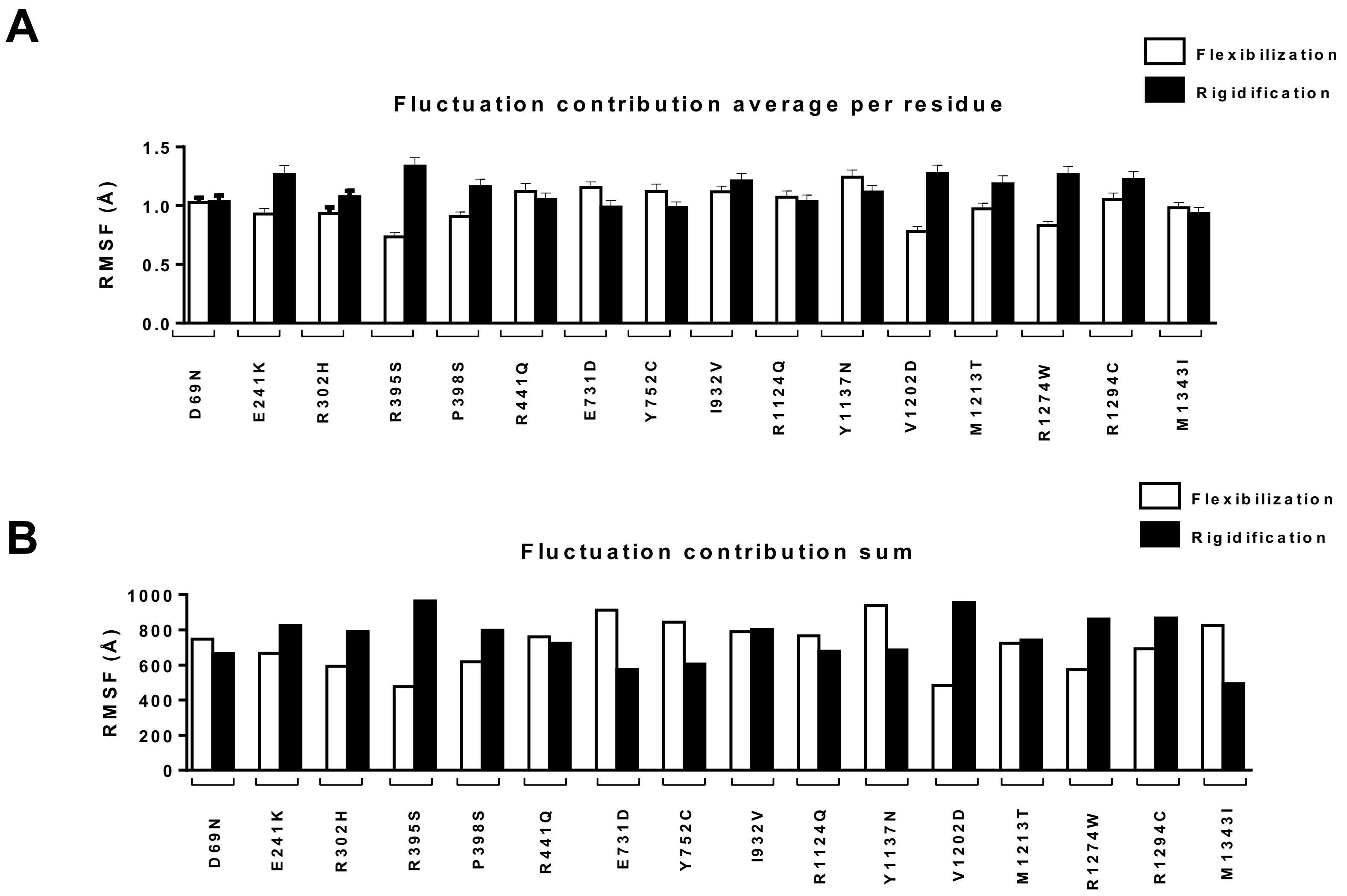


**Figure S4: Coarse-Grained molecular dynamics run.** Assessment of the flexibility contribution across the entire length of the protein, calculated by comparing the difference of RMSF of the WT minus RMSF of the mutant. Contribution to flexibility is shown in white bars, contribution to rigidity in black bars **A.** Average RMSF per residue for both contributing factors. **B.** Sum of RMSF values for both contributing factors.

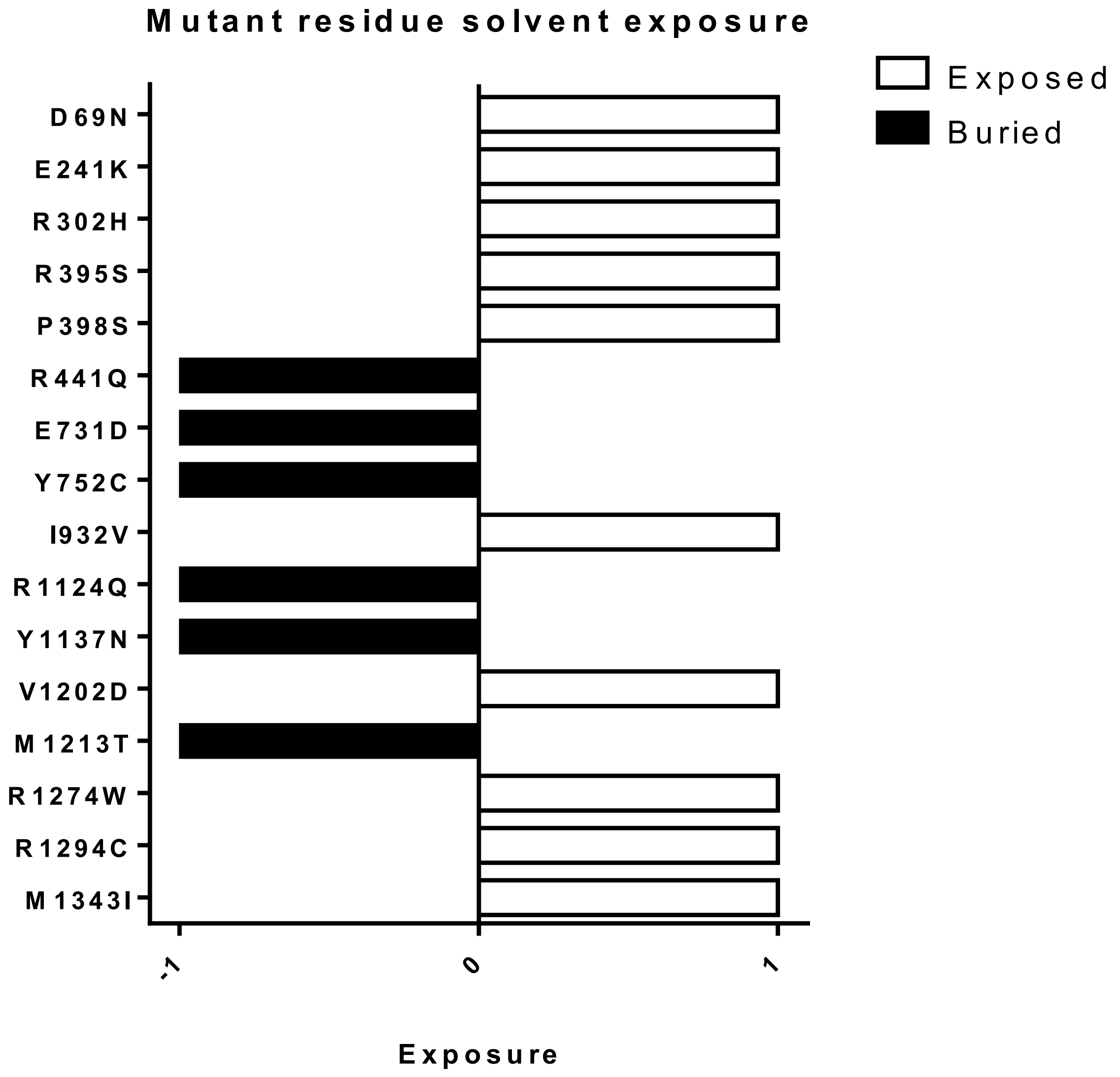


**Figure S5: Difference in solvent accessibility of mutant residues.** Exposure is expressed as -1 (black bars) for buried and +1 (white bars) for exposed residues. Six residues are buried, while 10 are exposed.

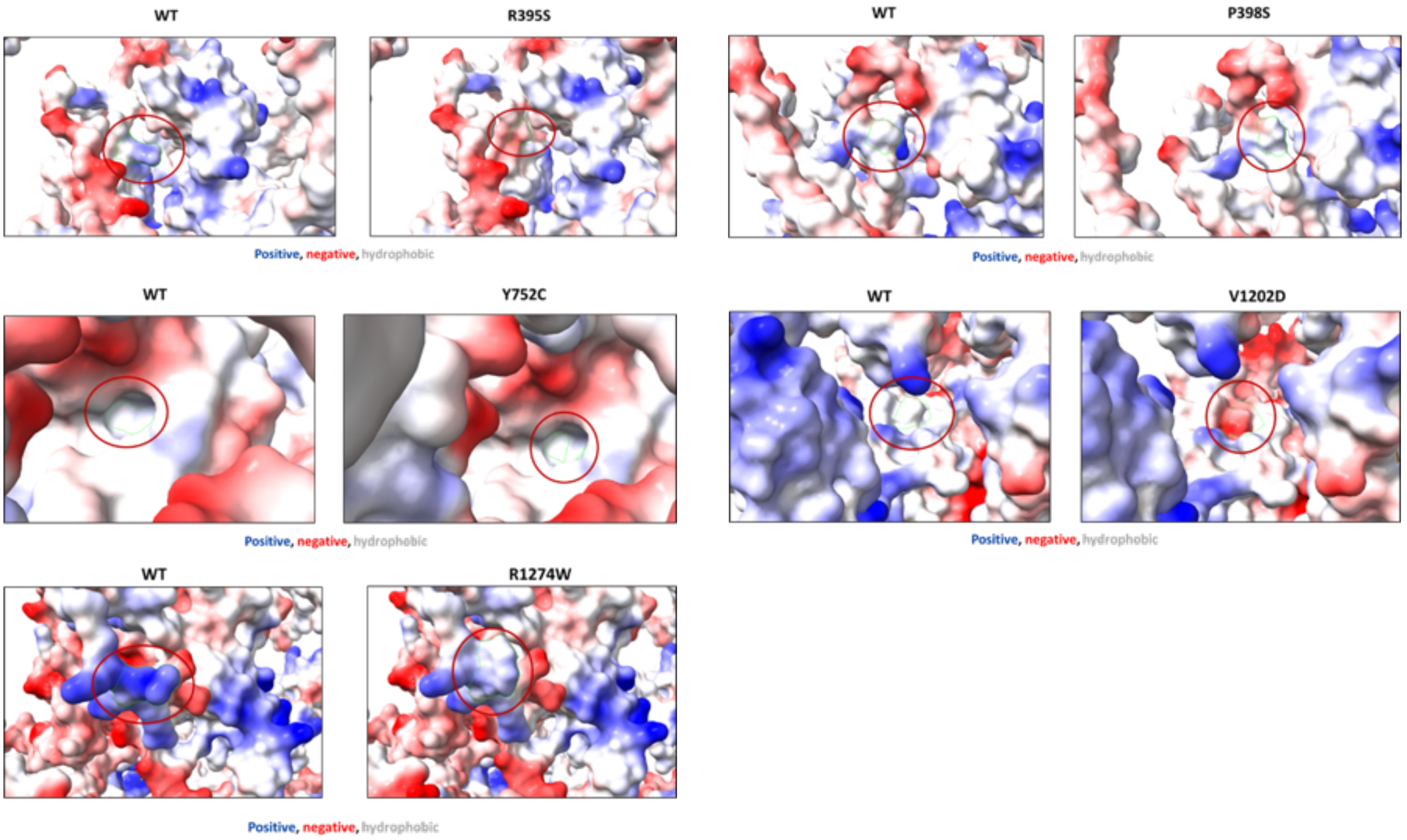


**Figure S6.** Surface analysis of several selected substitutions. Note that R1274W is predicted to have significant effects.

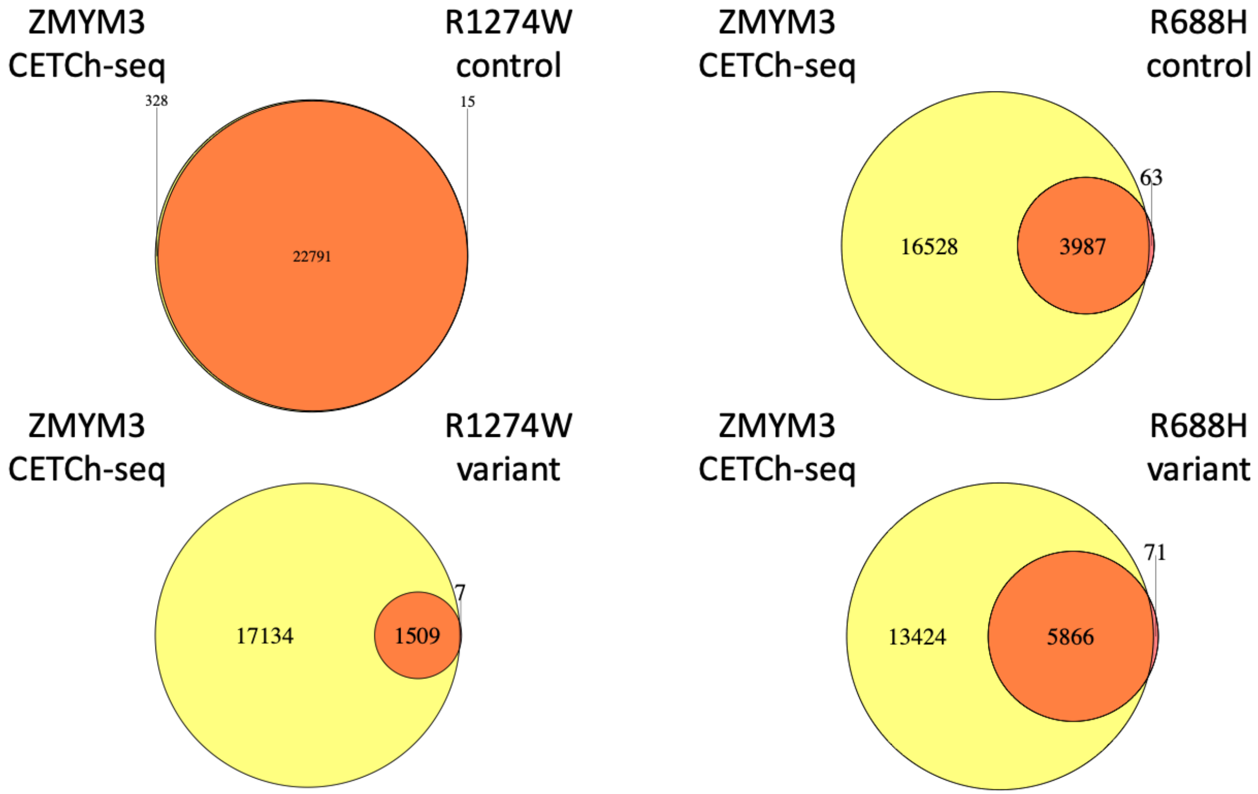


**Figure S7. Intersection of csaw analysis and peak calls.** For each pairwise comparison, the union of peaks was determined (downsampled to 20 million reads per replicate, IDR 0.05, merged peaks) and intersected with all csaw analyzed regions. Regions with significantly higher reads in the ZMYM3 CETCh-seq experiment are colored in yellow; regions with significantly higher reads in the comparison experiment are colored in red; and regions that are not significantly different are colored in orange (the overlap set; FDR cutoff for significance is 0.05). R1274W control is highly similar to ZMYM3 CETCh-seq, with the vast majority of regions not significantly differential. The other compared experiments are subsets of the ZMYM3 CETCh-seq experiment with very few differentially higher regions.

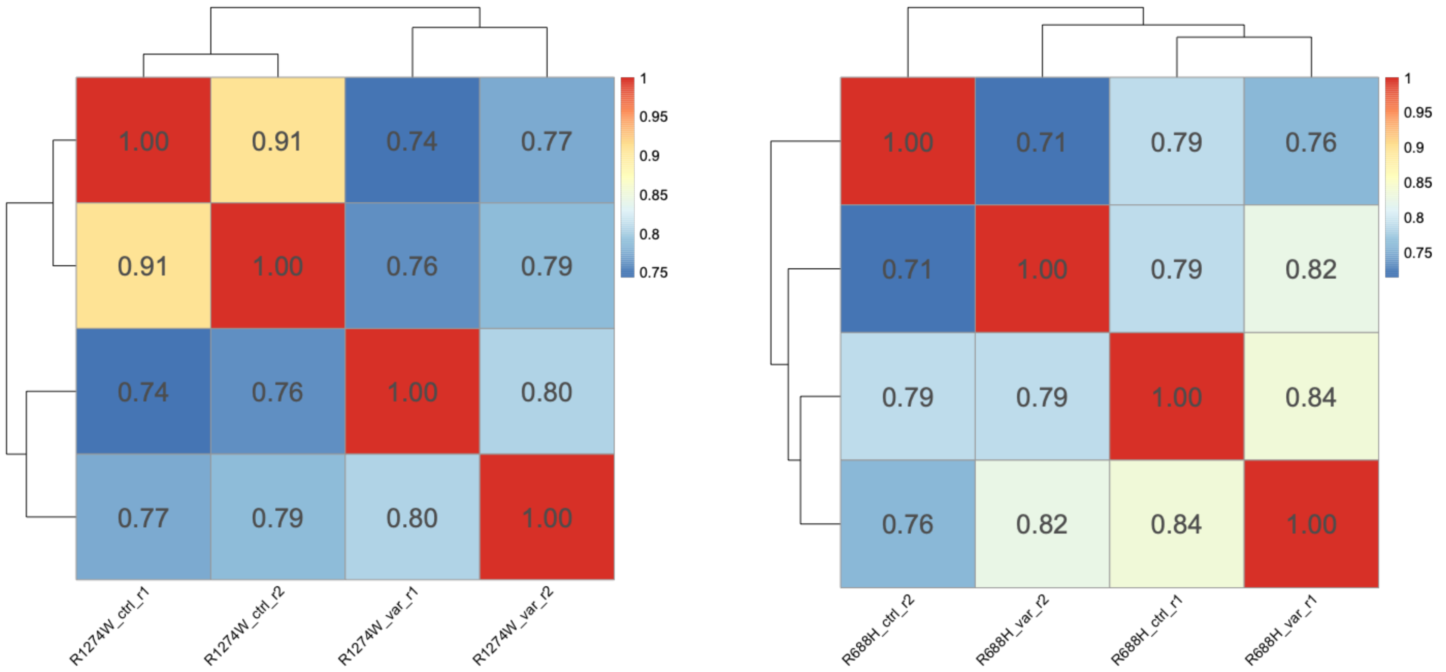


**Figure S8. Read count correlations.** The union of peaks between all experiments ($ZMYM3^{CETCh}$, $ZMYM3^{R1274W}$, and $ZMYM3^{R688H}$) was calculated, and reads from the .bam file for each replicate (downsampled to 20 million reads) were determined at each of these genomic positions. These data were correlated for each pairwise comparison and Pearson correlation coefficient was determined and plotted with clustering by pheatmap v. 1.0.12 in R v. 4.2.1.

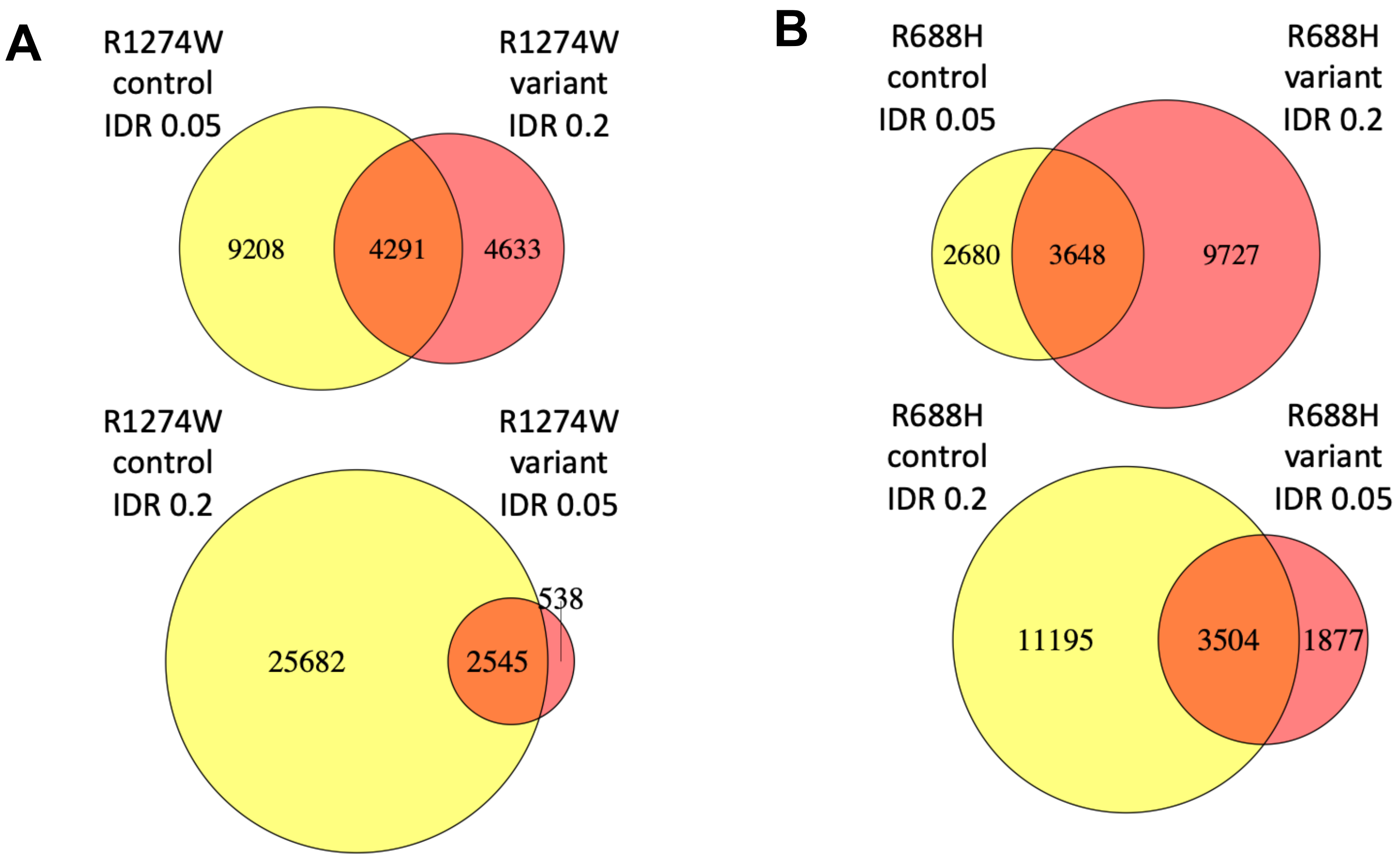


**Figure S9. Standard peak overlaps with relaxed peaks.** Peaks were called between replicate experiments downsampled to 20 million reads at the standard IDR threshold of 0.05 and also at a relaxed IDR threshold of 0.2. This analysis attempts to find peak overlaps missed by comparing standard peak calls due to enriched regions near peak-calling threshold. For R1274W (A), many regions in the variant experiment called as peaks in control are just under threshold and found by this approach, indicating a general reduced binding by the variant. For R688H (B), an experiment with greater noise, peaks are gained in both comparisons.
