## SupplementalMaterialsMethods for "Deleterious, protein-altering variants in the X-linked transcriptional coregulator *ZMYM3* in 22 individuals with a neurodevelopmental delay phenotype"

**Supplemental Materials and Methods**

**SEQUENCING**

Several probands had either exome sequencing (ES) or the Autism/ID Xpanded Panel (AIDX) through GeneDx, and details are listed in the table below. Using genomic DNA from the proband and parents (if submitted), the exonic regions and flanking splice junctions of the genome were captured using the IDT xGen Exome Research Panel v1.0 (Integrated DNA Technologies, Coralville, IA). Massively parallel (NextGen) sequencing was done on an Illumina system with 100 bp or greater paired-end reads. Reads were aligned to human genome build GRCh37/UCSC hg19 and analyzed for sequence variants using a custom-developed analysis tool. Reported variants were confirmed, if necessary, by an appropriate orthogonal method in the proband and, if submitted, in selected relatives. Additional sequencing technology and variant interpretation protocol has been previously described[1]. The general assertion criteria for variant classification are publicly available on the GeneDx ClinVar submission page (http://www.ncbi.nlm.nih.gov/clinvar/submitters/26957/).

| **Individual** | **Protein Effect** | **Testing details** |
|---|---|---|
| 1 | p.(D69N) | AIDX trio |
| 2 | p.(E241K) | ES duo with mom |
| 5 | p.(P398S) | AIDX trio |
| 6 | p.(R441Q) | ES trio |
| 15 | p.(R1274W) | AIDX duo with mom |
| 18 | p.(L226WfsX8) | ES trio |
| 19 | p.(Y752C) | AIDX trio, Sanger confirmation for proband but parents had high quality NGS data and Sanger confirmation was not necessary |

**Individual 3, R302H and Individual 7, R441Q**

Exome sequencing was performed on Individuals 3 and 7 as described[2]. The patients were enrolled for trio-WES as part of the international network of Autism Sequencing Consortium (ASC) (https://asc.broadinstitute.org/). Exome analysis (WES) was performed at the Broad Institute on Illumina HiSeq sequencers[3]. WES raw data of the trios were processed and analyzed using an in-house implemented pipeline[4,5].

**Individuals 4a, 4b, R395S**

Genomic DNA extracted from leukocytes of both patients and their parents was used for whole-exome sequencing. Exome enrichment was performed on individually barcoded samples using SeqCap EZ Exome Probes v3.0 (Roche) and sequencing was performed on HiSeq 2500 (Illumina) with 100bp paired-end reads. Reads were aligned to the hg19 reference genome using Novoalign version 3.02.13 (Novocraft) with default parameters.

After genome alignment, conversion of SAM format to BAM and duplicate removal was performed using Picard Tools (2.20.8). The Genome Analysis Toolkit, GATK (3.8)[6] was used for local realignment around indels, base recalibration, variant recalibration, and variant calling. Variants were annotated using the GEMINI framework[7] and filtered based on the population frequencies using several public databases and an in-house database of population-specific variants. Identification of candidate variants was performed for autosomal dominant (*de novo* variants) and autosomal recessive inheritance patterns. Variants were further prioritized according to the functional impact and conservation score. Sanger sequencing confirmed presence of the variants in the patients.

**Individual 8, R441Q**
Trio exome sequencing was performed for Proband 8. DNA was enriched using Agilent SureSelect Clinical Research Exome V2 capture and paired-end sequencing using the Illumina platform (outsourced). The aim was to obtain 8.1 Giga base pairs per exome with a mapped fraction of 0.99. The average coverage of the exome was ~50x. Duplicate reads were excluded. Data were demultiplexed with bcl2fastq Conversion Software from Illumina. Reads were mapped to the genome using the BWA-MEM, and variants were called using GATK HaplotypeCaller. Detected variants were filtered and annotated with Cartagenia software and classified with Alamut Visual.

**Individuals 9a, 9b, E731D**
WES was performed on gDNA of both Individuals 9a and 9b. The exome was captured using the xGen Exome Research Panel v2 (Integrated DNA Technologies) and sequenced using the Illumina HiSeq4000 platform according to the manufacturer's protocols. The overall mean-depth base coverage was 136- and 125-fold, while on average 93% and 92% of the targeted region was covered at least 20-fold, respectively for V.2 and V.3. Read mapping and variant calling were performed as described[8] using the Varapp software[9]. Sanger sequencing confirmed the segregation of the potentially causative variant.

**Individual 10, I932V**
Proband 10 had exome sequencing, and both short-read and long-read genome sequencing as described[10]. The variant was first observed in ES.

**Individual 11, R1124Q**
For Individual 11, patient blood was drawn into an EDTA blood collection tube. Isolation of DNA from whole blood was performed using the QIAsymphony (Qiagen). Sequencing libraries were constructed from patient whole blood genomic DNA using the HudsonAlpha Clinical Sequencing Lab's custom whole genome library preparation protocol. Patient DNA was sequenced on the Illumina HiSeqX sequencer. DNA library fragments were sequenced from both ends (paired) with a read length of 150 base pairs. Patient genomes were sequenced at an approximate depth of 30X, with at least 80% of base positions reaching 20X coverage. Sequence variants were called using GATK3 and loaded into a custom software analysis application for interpretation. All sequence variants were annotated with relevant information from established data

sources to provide support for variant interpretation. Variant pathogenicity was determined using ACMG criteria[11].

**Individual 12, Y1137N**

For Individual 12, Genomic DNA was extracted from EDTA blood of the patient and his parents. Whole Exome Sequencing (WES) on the patient was performed using the xGen® Exome Research Panel v1.0 (IDT) with paired-end sequencing (HiSeq SBS Kit v4, 125 Fwd-125 Rev, Q30-value: 84) on a HiSeq System (Illumina Inc.). Raw fastQ files were aligned to the hg19 reference genome using NextGene (Softgenetics). The average depth of coverage was 225x and 99.4% of the targeted bases were assessed by ≥20 independent sequence reads. By applying filters for known and candidate ID genes (SYSID and In-House) and Minor Allele Frequency ≤ 2% (gnomAD, ExAC) a total of 37 variants were observed in at least 16% of reads with sufficient quality level. Variants were investigated computationally for deleterious effects, by associations of the affected gene with proband's phenotype and by literature search for functional information. The candidate *ZMYM3* mutation from the WES approach was re-sequenced in the index, his mother and maternal grandparents after PCR amplification by Sanger sequencing using an ABI Genetic Analyzer 3730 (Applied Biosystems, Foster City, California.

**Individual 13, V1202D**

The proband and his parents had ES as described[12,13]. Parentage was confirmed with ES data and *de novo* status of the *ZMYM3* variant was confirmed by Sanger sequencing.

**Individual 14, M1213T**

A clinical comprehensive intellectual disability panel (555 genes) was ordered through Fulgent Genetics, which returned the *ZMYM3* variant here. Follow-up reanalysis through the Care4Rare project[14,15] also identified this *ZMYM3* variant as the top hit.

**Individual 16, R1294C**

Individual 16 had both trio exome sequencing and genome sequencing. For exome sequencing, DNA was extracted from blood samples using the Hamilton automate machine. Exome DNA library was prepared with the Agilent Focused Exome preparation kit. High-throughput sequencing was performed on a NextSeq5500 sequencer (Illumina) with a 2x75 bp paired-end running method. The BWA-MEM algorithm was used to map the reads on the reference genome (GRCh37/hg19). The variant calling was performed according to GATK and FreeBayes best practices. The ANNOVAR and ALAMUT (Interactive Biosoftware) tools were used for variant annotation. Genome sequencing was performed on the SeqOIA national platform.

**Individual 17, M1343I**

For Individual 17, a CHOP Medical Exome was performed on the proband and mother. Genomic DNA was extracted from peripheral blood or other patient tissues following standard DNA extraction protocols. After extraction of genomic DNA, targeted exons are captured with the Agilent SureSelect XT Clinical Research Exome kit (per manufacturer's protocol) and sequenced on the Illumina HiSeq 2000 or 2500 platform with 100bp paired-end reads. Mapping and analysis were based on the human genome build UCSC hg19 reference sequence. Sequencing data is processed using an in-house custom-built bioinformatics pipeline. The bioinformatics protocol utilized for this evaluation is version CWES-2.2. The exome sequencing protocol utilized for this evaluation is version 3.1. Coding exons and splice sites targeted with the exome kit are analyzed and reported. The following pathogenic variants are detectable: single nucleotide variants, small deletions and small insertions.

**Individual 20, R1294C**
Individual 20 had exome sequencing. ES was performed using Nimblegen SeqCap Ez MedExome Target Enrichment Kit (Roche Sequencing Solutions, Pleasanton, CA, USA) and an Illumina NextSeq500 (Illumina Inc., CA, USA) as paired-end 150 bp reads. Sequences were analyzed with the SeqOne platform (Montpellier, France). Sanger sequencing was performed for variant confirmation and segregation. X-chromosome inactivation study was performed by methylation analyses using the HUMARA assay[16].

**R1294C Cases**
We note that R1294C has been observed in two individuals here (probands 16 and 20), each confirmed to be *de novo*. Each of these cases were submitted by sites in Europe, and we note that a ClinVar submission (SCV000297052.2) was made by the Children's Hospital of Philadelphia, USA. While this does appear to be a unique case from those described here, we cannot confirm this.

### COMPUTATIONAL MODELING

The wild-type 3D protein structure was downloaded from AlphaFoldDB (https://alphafold.ebi.ac.uk/)[17], which was included with the reference from UniProt (Accession number: Q14202). When not possible online, structures were visualized, colored and the sequence was mutated with Chimera version 1.15, rotamer builder tool[18]. Specifically, structure superposition was obtained in Chimera with the tool Matchmaker. Structure refinement was performed with the Chimera tool Dock Prep with standard settings, as previously described[19]. Depiction of molecular surfaces was defined as VdW surface and colored according to the electrostatic potential.
**Wild-Type Protein model analysis**: The distinction between organized and disordered regions was based upon the uniprot reference. The pLDDT value for each residue was extracted from the pdb file (opened as text file) from the B-factor field as prescribed on https://alphafold.ebi.ac.uk/about. The PAE matrix was downloaded from the main page of AlphaFoldDB and analyzed with the web-interface built-in tool.
**Free folding energy estimation:** the ΔΔG was calculated according to the SimBaNI model developed by Caldarau and colleagues[20]. For the SASA term, the RSA obtained with FreeSASA and naccess parameters were considered. Variations >1.0 kcal/mol in module were considered as significant.
**Flexibility inspection:** The flexibility analysis was performed by submission of pdb structures to CABS-flex2 with no restraints[21]. The results, expressed as RMSF were downloaded and a cutoff of 1 Å was considered as relevant.

**Normal mode analysis:** The normal mode analysis was performed by submission of the WT pdb file to WebNMA[22] and run in a comparative mode by separately setting the mutation parameters. The deformation energy per-residue was then downloaded and plotted. We applied a cutoff of 2 kcal/mol.
**Surfaces calculation:** the molecular and solvent accessible surfaces were calculated with two methods. First, both molecular (VdW radii) and solvent-accessible (SASA, probe radius 1.4 Å) total, polar and (by difference) non-polar surfaces were computed to calculate the variations. To this aim, VegaZZ suite was employed[23]. Then, relative-solvent-accessible surface area (RSA) was calculated by submitting the pdb files to FreeSASA with the parameters derived from naccess[24]. Burial-exposure mutant residue classification: the classification into buried/exposed mutant residues was based on the computed RSA values. By comparing the RSAs with the tabulated values[25] and selecting a cutoff of 0.20.

### CHIP-SEQ EXPERIMENTS

We edited the genomic DNA at the *ZMYM3* endogenous locus in HepG2 cells to introduce the variant of interest simultaneously with a 3X FLAG tag, 2A self-cleaving peptide, and neomycin resistance gene, using a modified version of the previously published CRISPR epitope tagging ChIP-seq (CETCh-seq) protocol[26]. HepG2 cells were sourced from ATCC (HB-8065) and cultured using the recommended protocol. We identified a CRISPR/Cas9 sgRNA targeting a DNA cleavage site near each variant (Table S5) using established methods and cloned this sgRNA into pX330-U6-Chimeric_BB-CBh-hSpCas9 (Addgene plasmid #42230)[27]. We designed homology-directed repair donor templates composed of 400 bp of genomic sequence upstream (relative to coding direction of *ZMYM3*) of the cleavage site, exonic sequence surrounding the variant of interest, all exons downstream of the exon harboring the variant without the stop codon, the in-frame FLAG/P2A/NeoR cassette, and 400 bp of genomic sequence downstream of the cleavage site (Table S5). For control experiments, we repaired the genomic DNA with reference sequence at the variant position; for variant experiments, we used the variant nucleotide. For both experiments, an additional mutation was inserted to abolish the PAM site and block cleavage of edited sequence by Cas9. This mutation was a synonymous substitution for PAM sites in coding sequence. The donor templates were synthesized and cloned into the BamHI site of pUC19 by GenScript (Piscataway, NJ, USA). Genomic coordinates for the variant positions, the PAM site mutation positions, and the exons included in the super-exon for each experiment are shown in the table below.

| Variant | hg38 substitution | super-exon | PAM mutation |
|---|---|---|---|
| R441W | chrX:71,249,610 G>A | chrX:71,249,461-71,249,679<br>chrX:71,249,020-71,249,169<br>chrX:71,248,686-71,248,799<br>chrX:71,248,438-71,248,524<br>chrX:71,248,163-71,248,312<br>chrX:71,247,734-71,247,904<br>chrX:71,247,346-71,247,510 | chrX:71,249,611 G>A |

| | | | |
|---|---|---|---|
| | | chrX:71,246,595-71,246,690<br>chrX:71,246,354-71,246,512<br>chrX:71,245,986-71,246,098<br>chrX:71,245,669-71,245,842<br>chrX:71,245,340-71,245,483<br>chrX:71,244,790-71,244,891<br>chrX:71,244,303-71,244,470<br>chrX:71,243,829-71,243,978<br>chrX:71,242,971-71,243,084<br>chrX:71,242,171-71,242,422<br>chrX:71,241,229-71,241,342<br>chrX:71,240,919-71,241,107 | |
| R688H | chrX:71,247,819 C>T | chrX:71,247,734-71,247,904<br>chrX:71,247,346-71,247,510<br>chrX:71,246,595-71,246,690<br>chrX:71,246,354-71,246,512<br>chrX:71,245,986-71,246,098<br>chrX:71,245,669-71,245,842<br>chrX:71,245,340-71,245,483<br>chrX:71,244,790-71,244,891<br>chrX:71,244,303-71,244,470<br>chrX:71,243,829-71,243,978<br>chrX:71,242,971-71,243,084<br>chrX:71,242,171-71,242,422<br>chrX:71,241,229-71,241,342<br>chrX:71,240,919-71,241,107 | chrX:71,247,824 G>A |
| R1274W | chrX:71,241,327 G>A | chrX:71,241,229-71,241,342<br>chrX:71,240,919-71,241,107 | chrX:71,241,351 G>A |

For each of the control and variant experiments, we nucleofected two million HepG2 cells with 5 ug total plasmid DNA (2.5 ug sgRNA/Cas9 plasmid and 2.5 ug respective donor plasmid) using a Lonza Nucleofector Kit V with an Amaxa Nucleofector 2. Immediately after nucleofection, each experiment was split into two wells of a 6-well plate, and these two replicates were recovered and grown separately. Two days post-nucleofection, we began selection with Geneticin (Invitrogen 10131) at 300 ug/mL. Cells were grown under selection for two weeks, and afterwards continued to expand in non-selective media for another three to four weeks. The R441W experiment did not survive selection, and no further work was performed on this variant. Genomic DNA was purified from cells and used as template for PCR and Sanger sequence validation of edits using a Qiagen DNeasy Blood & Tissue Kit (Table S5). Cells (20 million for each replicate) were crosslinked and harvested, immunoprecipitation with M2 FLAG monocolonal antibody (Sigma F1804) was performed, and sequencing libraries were constructed, all as previously described[28]. The libraries were pooled with other CETCh-seq libraries and sequenced on a NovaSeq S2 flowcell yielding total aligned read counts as shown in Table S4. We performed peak calling using SPP[29] and Irreproducible Discovery Rate (IDR)[30] using ENCODE-standardized pipelines for analysis and quality-control[31]. We also downsampled all replicate bam files to 20 million reads and performed peak calling, using either the standard IDR cutoff of 0.05 or a relaxed IDR cutoff of 0.2. We performed additional differential binding analyses using the R package csaw v1.28.0[32], using window widths of 10 nucleotides and background bins of 10,000 nucleotides, and using downsampled (to 20 million reads) bam files for all replicates. We generated Activity-by-Contact (ABC) v0.2 loop calls[33] using hg38 reference sequence and gene coordinates downloaded from the UCSC genome table browser, the ENCODE datasets ENCSR149XIL (DNase-seq), ENCSR000AMO (H3K27Ac), and ENCFF356LFX (blacklist), RNA-seq for HepG2 downloaded from Expression Atlas, and processed HepG2 Hi-C data from 4D Nucleome (4DNESC2DEQIJ).

The overlaps of peaks called from downsampled bam files (20 million reads) were 67.8% between ZMYM3$^{R1274W}$-control and ZMYM3$^{R1274W}$-variant, and 46.9% between ZMYM3$^{R688H}$-control and ZMYM3$^{R688H}$-variant. Knowing that peak overlaps suffer from missed calls at regions near threshold in experiments, we expanded the peak overlap analysis to use peaks called at the standard IDR cutoff of 0.05 in one experiment and peaks called at a relaxed IDR cutoff of 0.2 in the other experiment. This increased overlaps to 82.5% and 65.1%, respectively (Figure S9). To examine experiment similarity agnostic to peak calls, we performed read count correlations using each separate replicate for each experiment. Rather than performing read count correlations across the entire genome (an analysis that suffers from the majority of regions being near background level of read counts), we filtered the analysis space to ZMYM3-specific regions. We determined the union of all peaks called in ZMYM3$^{CETCh}$, ZMYM3$^{R688H}$, and ZMYM3$^{R1274W}$, and used this set of regions for read counts. Reads from each bam file were determined at each genomic location and the Pearson correlation coefficient was calculated for each pairwise comparison (Figure S8). For R1274W, replicate experiments were highly correlated (r=0.80-0.91), and the correlation between control and variant experiments was high (r=0.74-0.79). For R688H, replicate experiments correlated similarly to correlations between control and variant (r=0.71-0.84). These results are consistent with the R688H experiments being of somewhat lower quality than R1274W experiments, and with no distinguishable difference between R688H control and variant beyond the noise of the assay.

**Supplemental Materials and Methods References:**


1. Retterer, K., Juusola, J., Cho, M.T., Vitazka, P., Millan, F., Gibellini, F., Vertino-Bell, A., Smaoui, N., Neidich, J., Monaghan, K.G., et al. (2016). Clinical application of whole-exome sequencing across clinical indications. Genet Med *18*, 696–704.
2. Pavinato, L., Trajkova, S., Grosso, E., Giorgio, E., Bruselles, A., Radio, F.C., Pippucci, T., Dimartino, P., Tartaglia, M., Petlichkovski, A., et al. (2021). Expanding the clinical phenotype of the ultra-rare Skraban-Deardorff syndrome: Two novel individuals with WDR26 loss-of-function variants and a literature review. Am. J. Med. Genet. A *185*, 1712–1720.
3. Satterstrom, F.K., Kosmicki, J.A., Wang, J., Breen, M.S., De Rubeis, S., An, J.Y., Peng, M., Collins, R., Grove, J., Klei, L., et al. (2020). Large-Scale Exome Sequencing Study Implicates Both Developmental and Functional Changes in the Neurobiology of

Autism. Cell *180*, 568-584.e23.
4. Bauer, C.K., Calligari, P., Radio, F.C., Caputo, V., Dentici, M.L., Falah, N., High, F., Pantaleoni, F., Barresi, S., Ciolfi, A., et al. (2018). Mutations in KCNK4 that Affect Gating Cause a Recognizable Neurodevelopmental Syndrome. Am. J. Hum. Genet. *103*, 621–630.
5. Flex, E., Martinelli, S., Van Dijck, A., Ciolfi, A., Cecchetti, S., Coluzzi, E., Pannone, L., Andreoli, C., Radio, F.C., Pizzi, S., et al. (2019). Aberrant Function of the C-Terminal Tail of HIST1H1E Accelerates Cellular Senescence and Causes Premature Aging. Am. J. Hum. Genet. *105*, 493–508.
6. McKenna, A., Hanna, M., Banks, E., Sivachenko, A., Cibulskis, K., Kernytsky, A., Garimella, K., Altshuler, D., Gabriel, S., Daly, M., et al. (2010). The Genome Analysis Toolkit: a MapReduce framework for analyzing next-generation DNA sequencing data. Genome Res *20*, 1297–1303.
7. Paila, U., Chapman, B.A., Kirchner, R., and Quinlan, A.R. (2013). GEMINI: integrative exploration of genetic variation and genome annotations. PLoS Comput Biol *9*, e1003153.
8. Alfaiz, A.A., Micale, L., Mandriani, B., Augello, B., Pellico, M.T., Chrast, J., Xenarios, I., Zelante, L., Merla, G., and Reymond, A. (2014). TBC1D7 Mutations are Associated with Intellectual Disability, Macrocrania, Patellar Dislocation, and Celiac Disease. Hum. Mutat. *35*, 447–451.
9. Delafontaine, J., Masselot, A., Liechti, R., Kuznetsov, D., Xenarios, I., and Pradervand, S. (2016). Varapp: A reactive web-application for variants filtering. BioRxiv 060806.
10. Cohen, A.S.A., Farrow, E.G., Abdelmoity, A.T., Alaimo, J.T., Amudhavalli, S.M., Anderson, J.T., Bansal, L., Bartik, L., Baybayan, P., Belden, B., et al. (2022). Genomic answers for children: Dynamic analyses of >1000 pediatric rare disease genomes. Genet. Med. *24*, 1336–1348.
11. Richards, S., Aziz, N., Bale, S., Bick, D., Das, S., Gastier-Foster, J., Grody, W.W., Hegde, M., Lyon, E., Spector, E., et al. (2015). Standards and guidelines for the interpretation of sequence variants: a joint consensus recommendation of the American College of Medical Genetics and Genomics and the Association for Molecular Pathology. Genet Med *17*, 405–424.
12. Motta, M., Fasano, G., Gredy, S., Brinkmann, J., Bonnard, A.A., Simsek-Kiper, P.O., Gulec, E.Y., Essaddam, L., Utine, G.E., Guarnetti Prandi, I., et al. (2021). SPRED2 loss-of-function causes a recessive Noonan syndrome-like phenotype. Am. J. Hum. Genet. *108*, 2112–2129.
13. Lin, Y.C., Niceta, M., Muto, V., Vona, B., Pagnamenta, A.T., Maroofian, R., Beetz, C., van Duyvenvoorde, H., Dentici, M.L., Lauffer, P., et al. (2021). SCUBE3 loss-of-function causes a recognizable recessive developmental disorder due to defective bone morphogenetic protein signaling. Am. J. Hum. Genet. *108*, 115–133.
14. Beaulieu, C.L., Majewski, J., Schwartzentruber, J., Samuels, M.E., Fernandez, B.A., Bernier, F.P., Brudno, M., Knoppers, B., Marcadier, J., Dyment, D., et al. (2014). FORGE Canada consortium: Outcomes of a 2-year national rare-disease gene-discovery project. Am. J. Hum. Genet. *94*, 809–817.
15. Hamilton, A., Tétreault, M., Dyment, D.A., Zou, R., Kernohan, K., Geraghty, M.T., FORGE Canada Consortium, Care4Rare Canada Consortium, Hartley, T., and Boycott,

K.M. (2016). Concordance between whole-exome sequencing and clinical Sanger sequencing: implications for patient care. Mol. Genet. Genomic Med. *4*, 504–512.
16. Bertelsen, B., Tümer, Z., and Ravn, K. (2011). Three new loci for determining x chromosome inactivation patterns. J. Mol. Diagn. *13*, 537–540.
17. Jumper, J., Evans, R., Pritzel, A., Green, T., Figurnov, M., Ronneberger, O., Tunyasuvunakool, K., Bates, R., Žídek, A., Potapenko, A., et al. (2021). Highly accurate protein structure prediction with AlphaFold. Nature *596*, 583–589.
18. Pettersen, E.F., Goddard, T.D., Huang, C.C., Couch, G.S., Greenblatt, D.M., Meng, E.C., and Ferrin, T.E. (2004). UCSF Chimera--a visualization system for exploratory research and analysis. J. Comput. Chem. *25*, 1605–1612.
19. Rossi Sebastiano, M., Ermondi, G., Hadano, S., and Caron, G. (2022). AI-based protein structure databases have the potential to accelerate rare diseases research: AlphaFoldDB and the case of IAHSP/Alsin. Drug Discov. Today *27*, 1652–1660.
20. Caldararu, O., Blundell, T.L., and Kepp, K.P. (2021). Three Simple Properties Explain Protein Stability Change upon Mutation. J. Chem. Inf. Model. *61*, 1981–1988.
21. Kuriata, A., Gierut, A.M., Oleniecki, T., Ciemny, M.P., Kolinski, A., Kurcinski, M., and Kmiecik, S. (2018). CABS-flex 2.0: a web server for fast simulations of flexibility of protein structures. Nucleic Acids Res. *46*, W338–W343.
22. Tiwari, S.P., Fuglebakk, E., Hollup, S.M., Skjærven, L., Cragnolini, T., Grindhaug, S.H., Tekle, K.M., and Reuter, N. (2014). WEBnm@ v2.0: Web server and services for comparing protein flexibility. BMC Bioinformatics *15*,.
23. Pedretti, A., Mazzolari, A., Gervasoni, S., Fumagalli, L., and Vistoli, G. (2021). The VEGA suite of programs: an versatile platform for cheminformatics and drug design projects. Bioinformatics *37*, 1174–1175.
24. Mitternacht, S. (2016). FreeSASA: An open source C library for solvent accessible surface area calculations. F1000Research *5*,.
25. Creighton, T.E. (2013). Proteins : structures and molecular properties (New York, NY, USA: Freeman).
26. Savic, D., Partridge, E.C., Newberry, K.M., Smith, S.B., Meadows, S.K., Roberts, B.S., Mackiewicz, M., Mendenhall, E.M., and Myers, R.M. (2015). CETCh-seq: CRISPR epitope tagging ChIP-seq of DNA-binding proteins. Genome Res. *25*, 1581.
27. Cong, L., Ran, F.A., Cox, D., Lin, S., Barretto, R., Habib, N., Hsu, P.D., Wu, X., Jiang, W., Marraffini, L.A., et al. (2013). Multiplex genome engineering using CRISPR/Cas systems. Science *339*, 819–823.
28. Meadows, S.K., Brandsmeier, L.A., Newberry, K.M., Betti, M.J., Nesmith, A.S., Mackiewicz, M., Partridge, E.C., Mendenhall, E.M., and Myers, R.M. (2020). Epitope tagging ChIP-seq of DNA binding proteins using CETCh-seq. Methods Mol. Biol. *2117*, 3–34.
29. Kharchenko, P. V., Tolstorukov, M.Y., and Park, P.J. (2008). Design and analysis of ChIP-seq experiments for DNA-binding proteins. Nat. Biotechnol. 2008 2612 *26*, 1351–1359.
30. Li, Q., Brown, J.B., Huang, H., and Bickel, P.J. (2011). Measuring reproducibility of high-throughput experiments. Https://Doi.Org/10.1214/11-AOAS466 *5*, 1752–1779.
31. Landt, S.G., Marinov, G.K., Kundaje, A., Kheradpour, P., Pauli, F., Batzoglou, S., Bernstein, B.E., Bickel, P., Brown, J.B., Cayting, P., et al. (2012). ChIP-seq guidelines and practices of the ENCODE and modENCODE consortia. Genome Res. *22*, 1813–

1831.
32. Lun, A.T.L., and Smyth, G.K. (2016). csaw: a Bioconductor package for differential binding analysis of ChIP-seq data using sliding windows. Nucleic Acids Res. *44*, e45.
33. Nasser, J., Bergman, D.T., Fulco, C.P., Guckelberger, P., Doughty, B.R., Patwardhan, T.A., Jones, T.R., Nguyen, T.H., Ulirsch, J.C., Lekschas, F., et al. (2021). Genome-wide enhancer maps link risk variants to disease genes. Nature *593*, 238–243.
